# DNA-guided CRISPR/Cas12 for RNA targeting

**DOI:** 10.1101/2024.11.21.24317744

**Authors:** Carlos Orosco, Santosh R. Rananaware, Boyu Huang, Michael P. Hanna, M. Reza Ahmadimashhadi, Jordan G. Lewis, Michael P. Baugh, August P. Bodin, Sarah J. Flannery, Ian H. Lange, Zoe R. Fang, Vedant N. Karalkar, Katelyn S. Meister, Piyush K. Jain

**Affiliations:** Department of Chemical Engineering, University of Florida, Gainesville, FL, USA; Department of Molecular Genetics and Microbiology, University of Florida, Gainesville, FL, USA; Department of Computer Science, University of Florida, Gainesville, FL, USA; Department of Chemical Engineering, Massachusetts Institute of Technology, Cambridge, MA, USA; UF Health Cancer Center, University of Florida, Gainesville, FL, USA

## Abstract

CRISPR-Cas nucleases are transforming genome editing, RNA editing, and diagnostics but have been limited to RNA-guided systems. We present ΨDNA, a DNA-based guide for Cas12 enzymes, engineered for specific and efficient RNA targeting. ΨDNA mimics a crRNA but with a reverse orientation, enabling stable Cas12-RNA assembly and activating trans-cleavage without RNA components. ΨDNAs are effective in sensing short and long RNAs and demonstrated 100% accuracy for detecting HCV RNA in clinical samples. We discovered that ΨDNAs can guide certain Cas12 enzymes for RNA targeting in cells, enhancing mRNA degradation via ribosome stalling and enabling multiplex knockdown of multiple RNA transcripts. This study establishes ΨDNA as a robust alternative to RNA guides, augmenting the potential of CRISPR-Cas12 for diagnostic applications and targeted RNA modulation in cellular environments.

## Main

Clustered Regularly Interspaced Short Palindromic Repeats (CRISPR) constitute an adaptive immune system in found prokaryotes, where CRISPR-associated (Cas) proteins complex with CRISPR RNAs (crRNAs) to precisely target and cleave exogenous nucleic acids ^1,2^. Since their discovery, CRISPR-Cas systems have been extensively developed for DNA and RNA genome editing and further engineered for applications such as diagnostics ^3–6^. Among the most well-known CRISPR-Cas effectors, type II (Cas9) and type V (Cas12) enzymes induce double-stranded breaks in DNA ^3,4,7^, while type VI (Cas13) targets and degrades RNA ^8,9^. Additionally, Cas12 and Cas9 exhibit non-specific collateral cleavage of single-stranded DNA (ssDNA) once the target sequence is bound, while Cas13 has similar activity against RNA. This functionality has been harnessed to develop various nucleic acid detection platforms ^5,6,10,11^. Overall, these advancements exploit the synergy between Cas effectors and crRNAs, enabling precise targeting of diverse nucleic acids.

As a quintessential part of the CRISPR system, crRNAs have been extensively studied to understand their interactions with Cas proteins ^1,2,12^. Transcribed from a CRISPR array, crRNAs are composed of direct repeats and spacer sequences. Once transcribed, the direct repeats are pre-processed to form a 5’ or 3’ handle, which constitutes the scaffold region of the crRNA ^7,13,14^. This scaffold region stabilizes the crRNA-Cas complex and aids in the recognition and binding of the target, while the adjacent spacer sequence is fully complementary to the target DNA or RNA. Given its unique structure and functionality, numerous efforts have been made to engineer crRNAs to enhance CRISPR-Cas systems. These efforts include incorporating modified bases ^15–17^, developing chimeric and split crRNAs ^18–22^, and altering their length ^23,24^. However, crRNA production is constrained by the complexity and expense of RNA synthesis, along with its shorter shelf life compared to DNA.

Naturally, CRISPR-Cas enzymes work in tandem with the crRNA to stabilize the complex and target a specific sequence ^3,4,7^. While there have been efforts to modify the target-binding spacer region of the guide RNA partially or completely with DNA bases for both Cas9 as well as Cas12, previous research has suggested that complete substitution of DNA bases within the guide RNA is typically not well-tolerated by both enzyme families ^25,26^. Contrary to this understanding, in this study, we demonstrated the ability of specific Type V systems (AsCas12a and Cas12i1) to withstand DNA guides for RNA targeting for cellular and *in vitro* applications. We engineered Cas12 enzymes, to adopt synthetic DNA mimics of a guide RNA (termed pseudo-guide DNA or ΨDNA) that assemble with Cas proteins, bind the target RNA and ultimately turn on trans-cleavage activity. We have successfully engineered ΨDNA guides to detect RNA substrates in a programmable manner.

To enable this discovery, we shortened the length of crRNA to only the spacer region while retaining the trans-cleavage activity of Type V CRISPR-Cas, specifically Cas12i1 and AsCas12a, against ssDNA targets. We then validated that a complementary DNA of the spacer region (cDNA) could turn on trans-cleavage activity in the presence of short RNA targets. Furthermore, the cDNA sequence was engineered with a 3’ DNA handle to mimic the structure of crRNAs (ΨDNA) to increase trans-cleavage activity for RNA targeting. Throughout the experiments, we observed that AsCas12a performed at a higher efficiency than Cas12i1 for inducing target-dependent trans-cleavage activity with ΨDNA guides. We further characterized how AsCas12a, ΨDNA, and target RNA bind together and ultimately developed a DNA-guided CRISPR/Cas-based RNA targeting platform. Finally, to demonstrate the applicability and adaptability of the ΨDNA-Cas12 complex, we validated its versatility by successfully testing Hepatitis C Virus-infected patient samples and evidenced its ability to induce translational repression in cells through ribosome stalling and subsequent mRNA degradation. This highlights the potential of ΨDNA-Cas12 technology as a cell-compatible tool for targeted RNA modulation within a cellular environment.

## Results

### Engineering of ΨDNA guides for RNA detection using Cas12 enzymes

To test the influence of crRNA scaffold length on the trans-cleavage activity of Cas12i1, we designed crRNAs with truncated scaffold sequences which include full-length crRNA, constructs with 5, 10, and 15 nucleotides removed from the 5’ end, and a spacer-only crRNA) (Fig. 1a, Supplementary Tables S1-2). The results showed Cas12i1 exhibited significant trans-cleavage activity with full-length crRNA and the minus 5 crRNA in the presence of double-stranded DNA (dsDNA) targets (Fig. 1b). Intriguingly for ssDNA targets, trans-cleavage activity was observed with the spacer-only crRNA. This suggests a unique scaffold-dependent interaction mechanism between the guide RNA and the target ssDNA or dsDNA. All trans-cleavage assays were performed on ssDNA reporters.

**Fig. 1:**
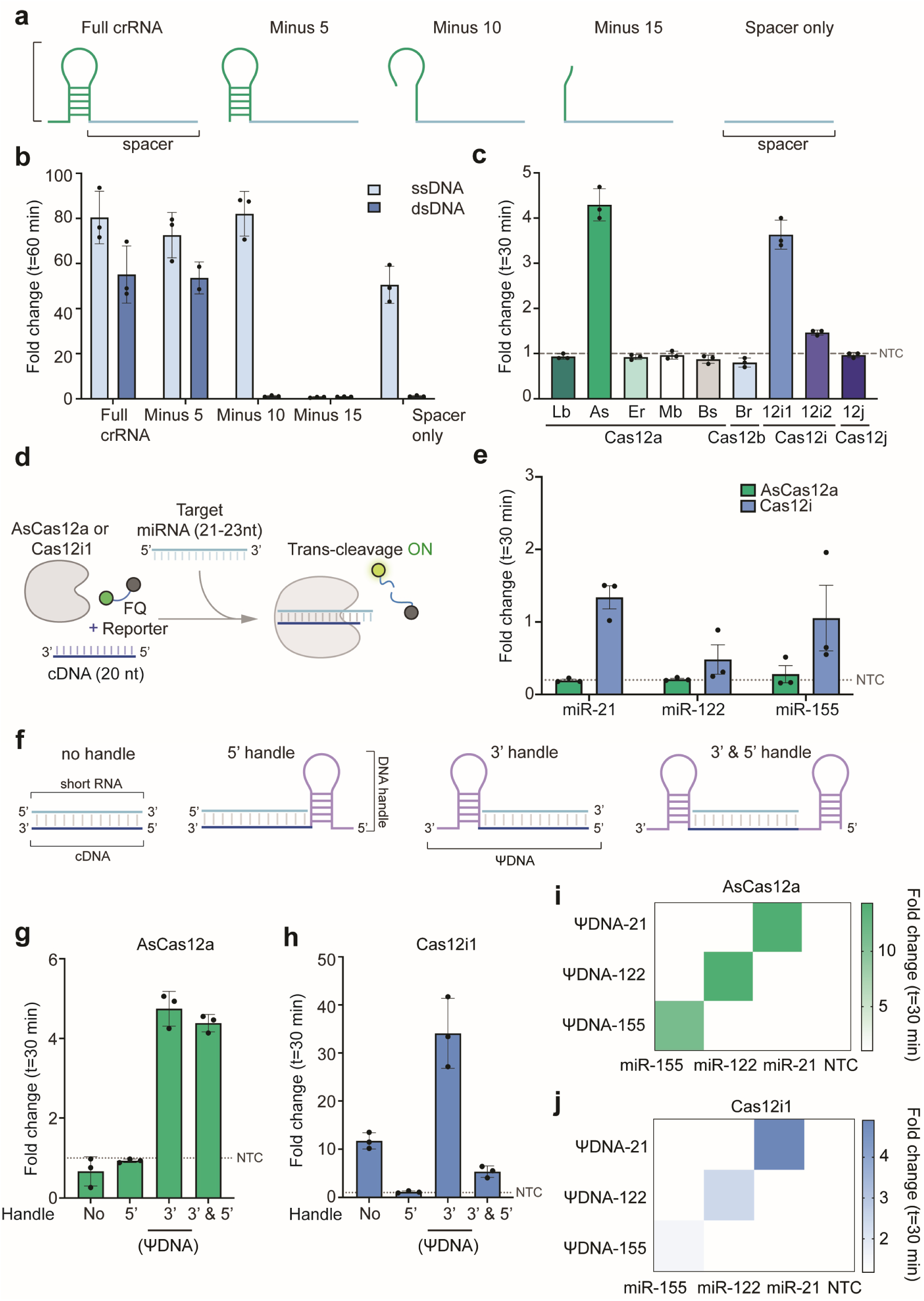
Engineering ΨDNA guides for RNA detection using Cas12 enzymes. **(a)** Schematic representation of different crRNA constructs used in this study: full-length crRNA with scaffold and spacer, and truncated crRNAs with 5, 10, and 15 nucleotides removed from the 5’ end of the scaffold, as well as a spacer-only crRNA lacking the scaffold. **(b)** Comparison of fold-change in trans-cleavage activity of Cas12i1 using different crRNA constructs in the presence of single-stranded DNA (ssDNA) and double-stranded DNA (dsDNA) targets. Full-length crRNA and minus 5 crRNA effectively induce trans-cleavage in the presence of dsDNA, whereas spacer-only crRNA induces activity only with ssDNA. **(c)** Fold-change compared to non-target (NTC) control in trans-cleavage activity of various Cas12 enzymes (LbCas12a, AsCas12a, ErCas12a, MbCas12a, BsCas12a, BrCas12b, Cas12i1, Cas12i2, and Cas12j) using spacer-only guide RNA with ssDNA targets. Cas12i1 and AsCas12a uniquely exhibit strong trans-cleavage activity with spacer-only guide RNA. **(d)** Schematic showing the mechanism of trans-cleavage activation by short c-DNA guides for miRNA detection. Cas12 enzymes complexed with c-DNA guides target miRNA, initiating trans-cleavage and subsequent fluorescence activation through the collateral cleavage of fluorescence-based reporters. **(e)** Fold-change compared to NTC control in the trans-cleavage activity of AsCas12a and Cas12i1 with c-DNA guides targeting miR-21, miR-122, and miR-155. Both enzymes show robust RNA detection with significant fold-change values for all three targets. **(f)** Schematic representation of different DNA guide designs: DNA without handle, with a 5’ handle, with a 3’ handle, and with both 3’ and 5’ handles. The handle in each case is a synthetic DNA mimic of the guide RNA scaffold. **(g-h)** Fold-change compared to NTC in trans-cleavage activity for AsCas12a and Cas12i1 using various DNA guide designs targeting miR-21. The 3’ handle DNA guide (termed ΨDNA) shows the highest detection activity for both enzymes. **(i-j)** Heatmap showing the specificity of ΨDNA guides (ΨDNA-21, ΨDNA-122, ΨDNA-155) for miRNA targets (miR-21, miR-122, miR-155) using AsCas12a and Cas12i1.

We extended our investigation to other Cas12 orthologs, including LbCas12a, AsCas12a, ErCas12a, MbCas12a, BsCas12a, BrCas12b, Cas12i2, and Cas12j, using the spacer-only guide RNA with ssDNA targets (Fig. 1c). Notably, Cas12i1 and AsCas12a displayed robust trans-cleavage activity, indicating that the ability to function with spacer-only guides may be specific to these enzymes.

Since a scaffold-independent spacer can be utilized for ssDNA detection, we tried to explore whether a ssDNA guide could be used for RNA detection. Firstly, we designed a 20-mer ssDNA with no scaffold (cDNA) and tested the system by targeting synthetic micro-RNAs (miRNA) (Fig. 1d). We assessed the fold-change in trans-cleavage activity of AsCas12a and Cas12i1 targeting three miRNA biomarkers: miR-21, miR-122, and miR-155. Cas12i1 demonstrated significant fold-change values for all three targets, underscoring the robust detection capability with cDNA guides (Fig. 1e). The difference between cDNA-guided detection for RNA targets and RNA spacer-guided detection for ssDNA targets is the concentration of ssDNA versus RNA.

To optimize guide DNA design and activate AsCas12a trans-cleavage, we experimented with various configurations of cDNA which include cDNA with a 5’ handle, cDNA with a 3’ handle, and cDNA with handles on both 5’ and 3’ ends (Fig. 1f). These handles are synthetic DNA mimics of the guide RNA scaffold. We found that the 3’ handle-modified cDNA guide, termed pseudo-DNA (ΨDNA), exhibited the highest detection activity for both AsCas12a and Cas12i1 when targeting miR-21, highlighting the importance of the handle’s position in enhancing detection efficiency (Figs. 1g-h). Comparing them, although Cas12i1 showed higher efficiency with the cDNA guide, AsCas12a performed better with the ΨDNA guide.

We further evaluated the specificity of ΨDNA guides for their respective miRNA targets to display the specificity of ΨDNA guides (ΨDNA-21, ΨDNA-122, ΨDNA-155) for miR-21, miR-122, and miR-155 with AsCas12a and Cas12i1, respectively. Each ΨDNA guide specifically detected its corresponding miRNA target with no off-target activity, demonstrating the high specificity of this approach (Figs. 1i-j). Additionally, 14 other miRNA sequences were detected successfully (Supplementary Fig. S1). To further characterize the ΨDNA trans-cleavage, we observed the ideal spacer length varies from 16 to 28 nucleotides (Supplementary Fig. S2), and short ΨDNAs are more sensitive to mismatches in the target sequences (Supplementary Fig. S3). Lastly, we observed RNA targeting with ΨDNA can withstand different divalent cations as catalysts for nucleic acid binding (Supplementary Fig. S4).

### Binding and efficiency of RNA cleavage by ΨDNA-Cas12 complexes

To understand and characterize the mechanism of ΨDNA-Cas12 complex formation, we conducted a series of experiments to compare the binding efficiency between ΨDNA and AsCas12a against its naturally occurring crRNA. First, we examined the assembly process of the AsCas12a-ΨDNA complex and its interaction with the target RNA through an Electrophoretic Mobility Shift Assay (EMSA). We compared the complex formation of AsCas12a with either a canonical crRNA guide targeting a single-stranded DNA (ssDNA) target, or a ΨDNA guide targeting an RNA target (Figs. 2a-b, Supplementary Table S3). The results demonstrate that ΨDNA guides form stable complexes with AsCas12a enzyme and effectively target RNA, similar to crRNA guides targeting ssDNA. Similar results were obtained for Cas12i1 and ΨDNA (Supplementary Fig. S5).

**Fig. 2:**
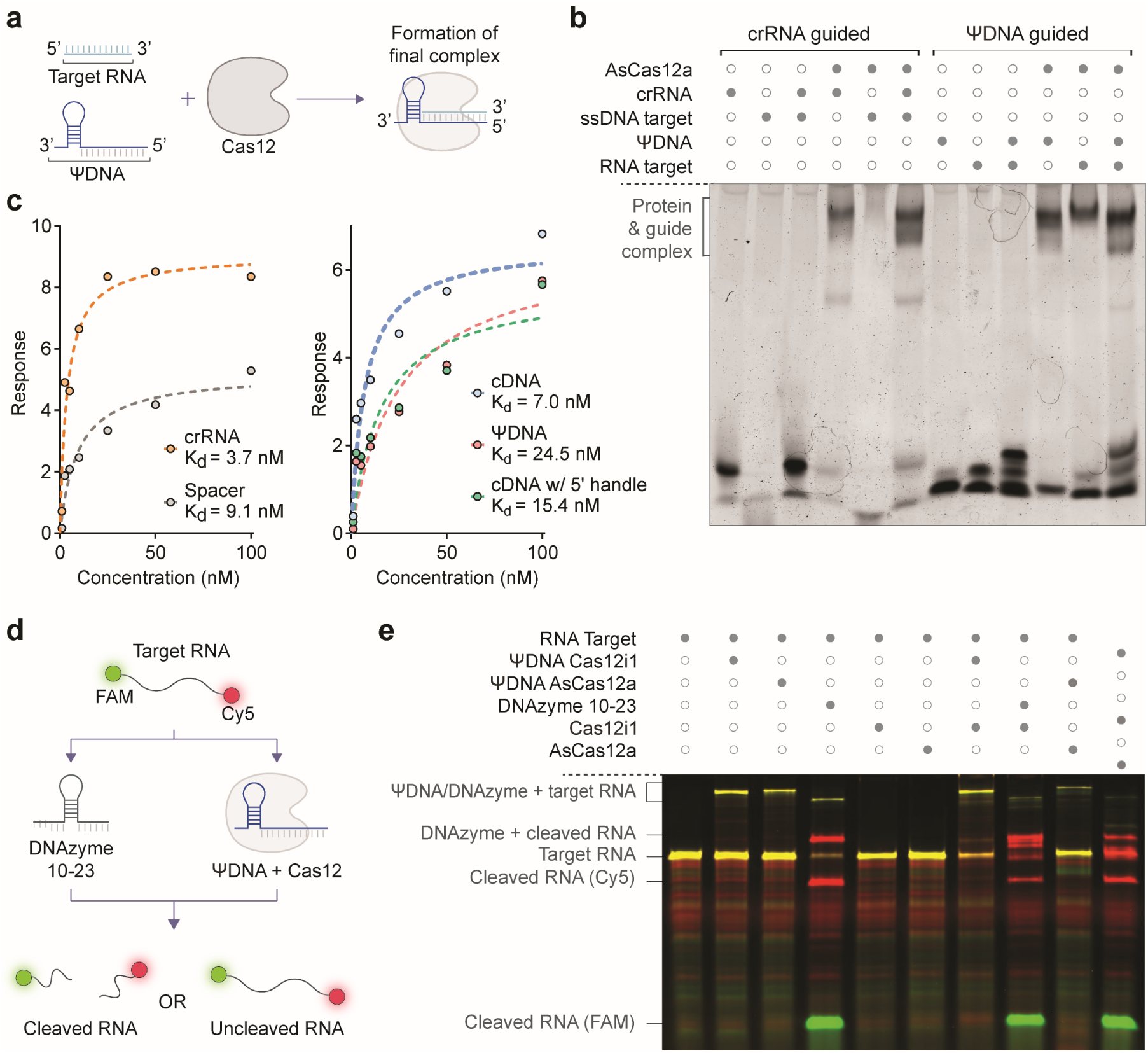
Binding and efficiency of RNA cleavage by ΨDNA-Cas12 complexes. **(a)** Schematic representation of the assembly process of the Cas12-ΨDNA complex and its interaction with the target RNA. The ΨDNA guide and Cas12 can bind to the RNA target with the 3’ handle of the ΨDNA occupying the space of the 5’ handle of the crRNA. **(b)** Electrophoretic Mobility Shift Assay (EMSA) comparing the complex formation of AsCas12a with a canonical crRNA guide targeting a ssDNA target (lanes 1-6) versus a ΨDNA guide targeting an RNA target (lanes 7-12). The dark circles indicate the presence of specific components in each lane. This gel demonstrates the ability of ΨDNA guides to form stable complexes with AsCas12 and target RNA. **(c)** Binding affinity (Kd) values for different guide constructs obtained via Bio-Layer Interferometry (BLI). This includes crRNA, spacer, cDNA, and cDNA with 5’ and 3’ handles (ΨDNA). **(d)** Schematic illustrating the mechanism of RNA cleavage by DNAzyme and ΨDNA-Cas12 complexes. The RNA target is labeled with FAM on the 5’-end and Cy5 on the 3’-end. The ΨDNA guide and DNAzyme induce cleavage of the RNA target, which can be detected by the separation of the FAM and Cy5 signals. **(e)** Gel electrophoresis showing the cleavage products of RNA targets treated with DNAzyme and ΨDNA-Cas12 complexes. The presence of cleaved RNA fragments is indicated by the separation of FAM and Cy5 signals. This gel demonstrates the effectiveness of ΨDNA-Cas12 complexes in cleaving RNA targets, with distinct bands indicating successful cleavage.

To further validate the observed interaction between ΨDNA and AsCas12a, we employed Bio-Layer Interferometry (BLI) assay to measure the dissociation constant (Kd) and quantify the binding affinity of various constructs. We tested the interactions between AsCas12a and several guides: crRNA, spacer (RNA), cDNA, cDNA with a 5’ handle, and ΨDNA (Fig. 2c, Supplementary Table S4). Similar to previous studies ^23^, the results revealed that AsCas12a exhibited the highest binding affinity for crRNA (Kd = 3.7 nM) and the difference among all DNA constructs and crRNA was less than 10-fold. Notably, ΨDNA demonstrated a dissociation constant within the low nanomolar range (Kd = 24.5 nM), indicating that the pseudo-guide binds efficiently to AsCas12a.

We next investigated the cis-cleavage activity of the ΨDNA-Cas12 complex to determine its ability to cleave RNA. To this end, we designed an RNA target labeled with FAM at the 5’-end and Cy5 at the 3’-end and used DNAzyme 10-23 as a positive control due to its known ability to cleave RNA ^27^ (Fig. 2d, Supplementary Table S5). After PAGE gel electrophoresis analysis, the results showed that the RNA target was cleaved only in the samples treated with DNAzyme 10-23, indicating that while ΨDNA-Cas12 complexes can trigger trans-cleavage activity in the presence of target RNA, they cannot perform cis-cleavage (Fig. 2e). Additionally, we observed no trans-cleavage on RNA with ΨDNA-mediated RNA targeting (Supplementary Fig. S6). Collectively, the data suggests that RNA binding to the complex triggers ΨDNA/Cas12a-mediated trans-cleavage activity only on ssDNA.

### Sensitivity of ΨDNA guides and scaffold sequence optimization for enhanced RNA detection using Cas12 enzymes

Having characterized the mechanism of ΨDNA, we then aimed to assess the applicability and specificity of ΨDNA guides for the detection of long RNA targets (∼1.7 kb) using Cas12 enzymes. Initially, synthetic gene fragments were utilized for *in vitro* transcription (IVT) to synthesize RNA, which were then used for Cas12-mediated detection with targeting and non-targeting ΨDNA guides. (Fig. 3a, Supplementary Table S6). We first evaluated this construct with the detection of synthetic viral HIV RNA fragments using AsCas12a and a library of 26 targeting and 22 non-targeting ΨDNA guides. The results illustrate a clear distinction between the signals from targeting ΨDNA guides, of which 92.3% (24 out of 26) showed a positive signal (Fig. 3b). However, only 34.6% (9 out of 26) of the targeting ΨDNA guides demonstrated high activity for Cas12i1 (Fig. 3c). Although both AsCas12a and Cas12i1 non-targeting ΨDNAs showed 100% accuracy, we concluded that AsCas12a has a higher flexibility for long RNA detection. Additionally, we successfully used ΨDNA-mediated detection against several endogenous mRNA targets (Supplementary Fig. S7).

**Fig. 3:**
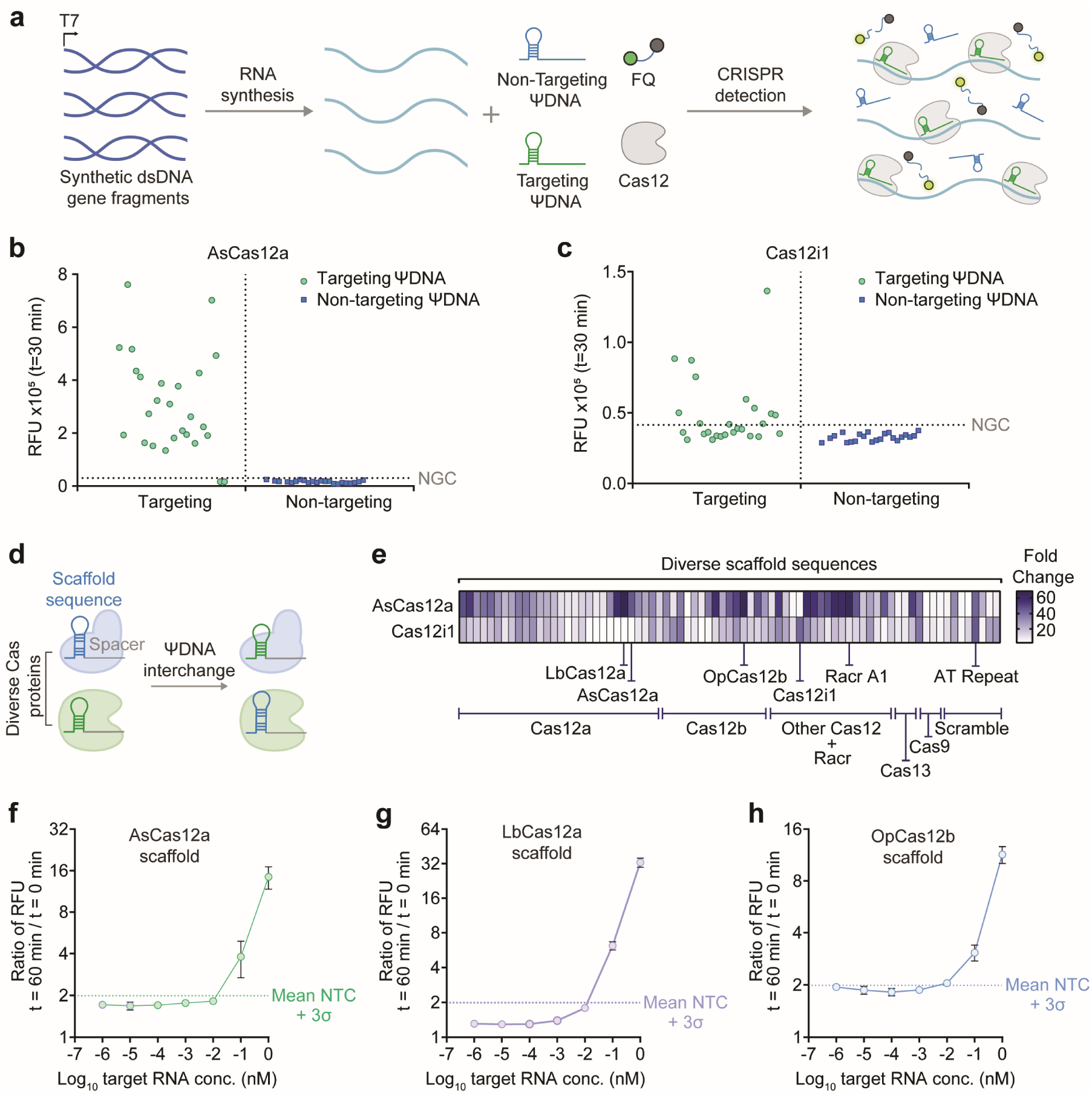
Application and specificity of ΨDNA guides in RNA detection using Cas12 enzymes. **(q)** Schematic representation of the experimental workflow for RNA detection using ΨDNA guides and Cas12 enzymes. Synthetic gene fragments are used for RNA synthesis via IVT, which are then combined with targeting and non-targeting ΨDNA guides. The Cas12 enzyme complexed with ΨDNA guides is employed for CRISPR detection, where the presence of target RNA activates trans-cleavage, leading to the collateral cleavage of ssDNA fluorescence-based reporters (FQ) which results in a detectable fluorescence signal. **(b)** Detection of synthetic HIV RNA fragments by AsCas12a using targeting and non-targeting ΨDNA guides. The relative fluorescence units (RFU) are plotted, showing a clear distinction between the signals from targeting ΨDNA guides (green dots) and non-targeting ΨDNA guides (blue dots). Targeting guides show significant fluorescence above the threshold, indicating successful detection of RNA targets. **(c)** Detection of synthetic HIV RNA fragments by Cas12i1 using targeting and non-targeting ΨDNA guides. Similar to AsCas12a, targeting ΨDNA guides (green dots) with Cas12i1 show higher fluorescence signals compared to non-targeting guides (blue dots), demonstrating effective RNA detection with lower efficiency than that of AsCas12a. **(d)** Schematic illustration of the interchange of scaffold sequences in different Cas enzymes to test the tolerance of scaffold mismatches and the impact on RNA detection efficiency. **(e)** Heatmap displaying the tolerance of AsCas12a and Cas12i1 to different direct repeat sequences derived from diverse Cas12 systems. The heatmap illustrates the relative fluorescence intensity, indicating the detection efficiency. Some non-canonical scaffold sequences enhance the detection activity compared to their canonical scaffolds, suggesting potential for further optimization of ΨDNA guides. **(f-h)** Sensitivity analysis of RNA detection using different scaffold sequences with AsCas12 enzyme. Ratio between RFU at *t* = 60 minutes versus t = 0 minutes at different target RNA concentrations for AsCas12a **(f)**, LbCas12a **(g)**, and OpCas12b **(h)** scaffold sequences. Error bars represent mean value +/- standard deviation (SD) (*n* = 3).

As previous discoveries have shown, switching the scaffold sequence of the crRNAs can enhance the activity of Cas12 enzymes ^11,28^. Thus, we explored optimizing RNA detection by varying the 3’handle sequence of ΨDNA by testing 77 different scaffold sequences derived from diverse CRISPR-Cas systems, including those from subtypes and orthologs of Type II, V, and VI (Fig. 3d, Supplementary Table S7). Using a fluorescence-based assay, we evaluated the detection efficiency of different scaffold sequences when paired with AsCas12a and Cas12i1 enzymes (Fig. 3e). The results illustrated variations in detection efficiency, revealing that some non-canonical scaffolds matched or exceeded the signal produced by the wild-type scaffold sequences. This enhancement suggests that specific sequences could be strategically chosen to optimize ΨDNA guides, thereby improving the performance of ΨDNA-based RNA detection. AsCas12a showed the highest performance with LbCas12a, OpCas12b, and its own scaffold. Cas12i1 showed the best activity with its wild-type scaffold sequence and with an AT repeat. Additionally, we observed higher flexibility of ΨDNA constructs with Cas12 enzymes as they can accept a wide variety of scaffolds for RNA detection compared to their crRNA counterparts for DNA targeting. (Supplementary Fig. S8).

To determine the sensitivity of RNA detection using different scaffold sequences, we conducted a sensitivity analysis with AsCas12a utilizing the scaffold sequences with the highest activity, these include sequences from AsCas12a, LbCas12a, and OpCas12b. By measuring the ratio of RFU at different target RNA concentrations we highlighted the limit of detection for AsCas12a. LbCas12a scaffold sequence exhibited the highest sensitivity (1-10 pM), with a lower limit of detection than the canonical AsCas12 scaffold underscoring the optimization of ΨDNA for RNA detection (Figs. 3f-h).

### Viral RNA detection and HCV patient sample validation with ΨDNA

To further demonstrate the capabilities of the Cas12-ΨDNA system for long RNA detection, we designed ΨDNAs to target various viral RNAs. As previously shown, the sensitivity of our construct ranges from 1-10 pM, necessitating a pre-amplification step for patient sample validation. We developed a two-pot reaction, where the viral RNA was pre-amplified via RT-PCR and then added to a T7 RNA polymerase and Cas12-ΨDNA reaction. This is similar to previous strategies used for Cas13 detection ^29^ (Fig. 4a). Using this construct, we successfully detected different viral targets, including HCV, Dengue, and Zika viral RNA (Fig. 4b, Supplementary Table S8). Specifically for the HCV virus, we targeted one conserved gene (5’UTR), and two highly mutated genes (NS5B and E2).

**Fig. 4:**
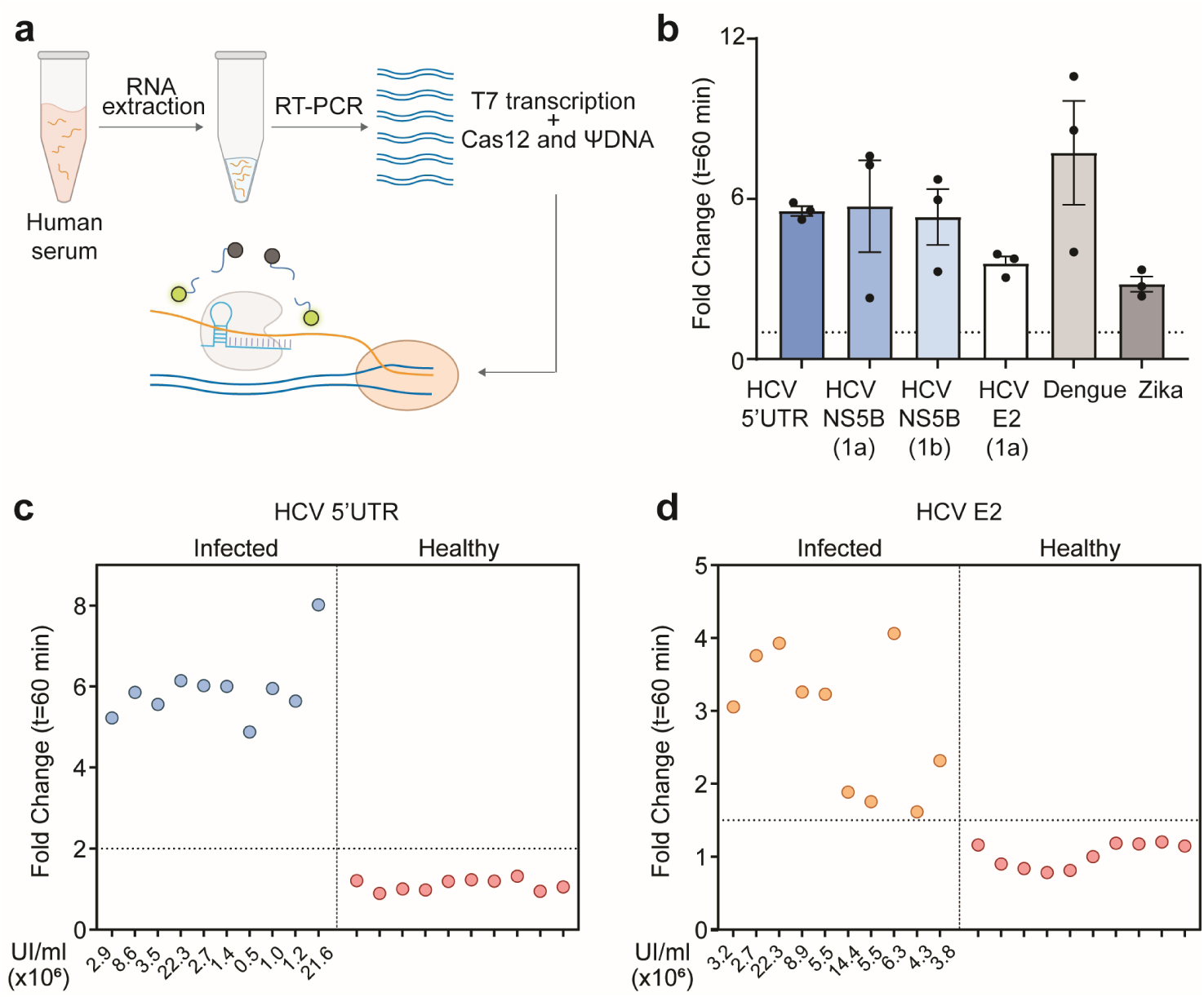
Binding and efficiency of RNA cleavage by ΨDNA-Cas12 complexes. **(a)** Schematic representation of viral RNA detection in patient samples. RNA is extracted from the sample followed by pre-amplification with RT-PCR. The amplified product is later added to a T7 transcriptase and ΨDNA-AsCas12a reaction mixture. **(b)** Fold-change compared to NTC in trans-cleavage activity for detection of synthetic mimics of viral RNA targets which include HCV (5’UTR, E2, and NS5B genes), Dengue, and Zika virus. Error bars represent mean value +/- standard deviation (SD) (*n* = 3). **(c-d)** Fold change in trans-cleavage activity for ΨDNA-AsCas12a mediated patient sample detection for HCV-positive and negative targets. A total of 40 samples were tested (20 for the 5’UTR gene and 20 for the E2 gene). The threshold was defined at a fold change equal to 2 for the 5’UTR gene and 1.5 for the E2 gene. For positive samples, UI/ml shown for viral load quantification.

We then validated our construct through HCV patient sample detection, successfully identifying 40 patient samples (20 positive and 20 negative) for two HCV genes (Figs. 4c-d). The positive patient samples tested with the 5’UTR ΨDNA included HCV-1a and HCV-1b phenotypes as it is a highly conserved gene ^30^. For the E2 gene, we designed the targeting-ΨDNA for the HCV-1a phenotype, as this region is highly mutated. Additionally, we performed NGS on samples to confirm the sequence and presence of viral HCV (Supplementary Fig. S9). Overall, we detected and discriminated all the samples with 100% accuracy.

### Translational repression in HEK293T cells with ΨDNA and AsCas12a

After characterizing the ΨDNA-Cas12 complex *in vitro*, we proceeded to evaluate its functionality in living cells, aiming for potential *in vivo* applications. We tested the ability of the DNA-guided AsCas12a complex to bind RNA within a cellular environment by targeting a specific mRNA sequence to induce ribosome stalling, thereby repressing translation and reducing protein synthesis. To achieve this, we designed a reporter system in which HEK293T cells were co-transfected with two plasmids: one encoding AsCas12a tagged with GFP and the other encoding mCherry. Additionally, we introduced ΨDNA constructs targeting the mCherry mRNA either at the start codon (ΨDNA1 and ΨDNA1+) or downstream (ΨDNA2), to observe the effect on mCherry translation repression (Fig. 5a). The ΨDNA constructs were modified with phosphorothioate ends to prevent exonuclease degradation within cells, and ΨDNA1+ included locked nucleic acids (LNAs) flanking the spacer region to enhance heteroduplex binding.

**Fig. 5:**
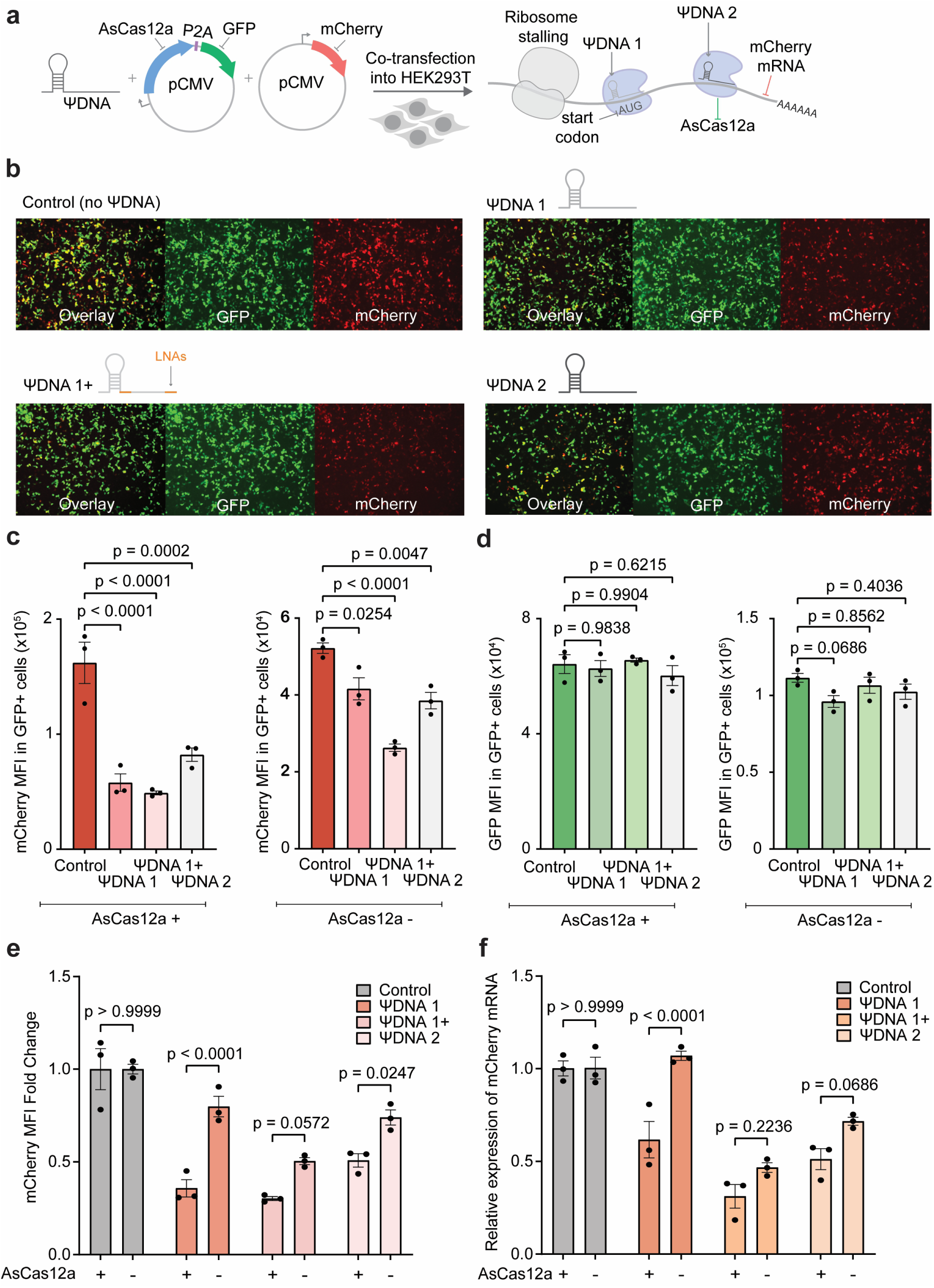
Translational repression of mCherry with ΨDNA-AsCas12a complex in HEK293T cells. **(a)** Schematic representation for co-transfection of AsCas12a-GFP, mCherry, and ΨDNA to induce ribosome stalling inside HEK293T cells. ΨDNA1 and ΨDNA1+ target the start codon of mCherry mRNA and ΨDNA2 targets the coding region. **(b)** Microscopy images for cells treated with AsCas12a-GFP, mCherry, and four different conditions of ΨDNA (no guide, ΨDNA1, ΨDNA1+, and ΨDNA2). Individual GFP and mCherry channels are shown as well as the overlay. **(c)** Geometric Mean Fluorescence Intensity (MFI) of mCherry for all four ΨDNA conditions with AsCas12a-GFP and GFP only. mCherry MFI is calculated only from GFP-positive cells. Geometric means are used for all calculations. **(d)** MFI of GFP on all GFP+ cells. The difference in expression of GFP is non-significant throughout all samples compared to the control. Similar GFP MFI demonstrates equal conditions for all samples. Geometric means are used for all calculations. (**c** & **d**) Statistical analysis for *n* = 3 biologically independent replicates were performed using Dunnet’s multiple comparison test against the control samples. Error bars represent the mean value +/− standard error of mean (SEM). **(e)** mCherry MFI fold change for all ΨDNA constructs with and without AsCas12a. Statistical analysis for *n* = 3 biologically independent replicates were performed using Sidaks’s multiple comparison test to compare the effect of AsCas12a on mCherry production. **(f)** Relative quantification of mCherry mRNA using 2^−ΔΔCt^ method. GAPDH was chosen as the endogenous control. Statistical analysis for *n* = 3 biologically independent replicates was performed using Sidaks’s multiple comparison test to compare the effect of AsCas12a on mCherry mRNA degradation. Each biological replicate had 3 technical replicates.

To assess the effects of ΨDNA and AsCas12a, we monitored the levels of GFP and mCherry 16 hours post-transfection using fluorescence microscopy (Fig. 5b, Supplementary Table S9). In our initial observations, the control group, which lacked ΨDNA, exhibited higher overall red fluorescence compared to the ΨDNA-treated samples. Among the treated groups, those with ΨDNA targeting the start codon showed a particularly noticeable reduction in red fluorescence intensity.

To further quantify the changes in mCherry production, we subjected the samples to flow cytometry. To ensure that any observed effects were not due to the ΨDNA alone triggering an antisense oligonucleotide (ASO) RNA degradation pathway, we included an additional control group that received GFP only, instead of the AsCas12a-GFP construct. Following flow cytometry, we compared the mean fluorescence intensity (MFI) of mCherry in all GFP-positive cells to determine whether the presence of AsCas12a and ΨDNA led to a reduction in overall red fluorescence intensity. The MFI of cells treated with AsCas12a and ΨDNA was significantly lower, confirming our initial observation that targeting the start codon is more effective. While samples treated with ΨDNA alone also showed reduced mCherry levels, the decrease was less pronounced compared to those treated with AsCas12a (Fig. 5c). To rule out any potential bias due to differences in transfection efficiency of AsCas12a-GFP or GFP alone, we confirmed that the MFI of GFP across all groups remained consistent (Fig. 5d). Comparing the fold change in mCherry MFI across the different ΨDNA constructs relative to the control clearly demonstrated that AsCas12a significantly contributed to mCherry translation repression. Although ΨDNA alone reduced mCherry fluorescence, particularly with ΨDNA1+, the presence of AsCas12a further enhanced this effect, leading to a significant decrease in protein expression across all constructs (Fig. 5e). Notably, ΨDNA1 exhibited the highest activity when paired with AsCas12a.

To assess RNA expression within the samples, we extracted total RNA and evaluated mRNA expression levels (Fig. 5f). In all samples, the presence of AsCas12a led to a noticeable decrease in mCherry mRNA levels. Given that the ΨDNA complex is incapable of RNA cis or trans-cleavage (Fig. 2e, Supplementary Fig. S6) we hypothesize that this mRNA degradation is induced by a no-go decay (NGD) mechanism ^31^. NGD is triggered when ribosomes stall and fail to elongate, leading to the degradation of mRNA that cannot produce functional proteins. Additionally, we observed that ΨDNA1+ alone triggered ASO-driven RNA degradation, as indicated by both MFI and mRNA levels, likely due to the strong hybridization induced by the incorporated LNAs. Nonetheless, AsCas12a significantly enhances gene repression, resulting in overall lower protein levels when both the protein and guide are present, confirming the effectiveness of the ΨDNA-Cas12a complex in HEK293T cells.

### Efficient endogenous RNA knockdown using ΨDNA-guided CRISPR-Cas12a

We assessed the ability of the ΨDNA-guided CRISPR-Cas12a system to selectively knockdown endogenous RNA transcripts in HEK 293T cells. To this end, we measured the relative expression levels of the PPIA, RPL4, and PCSK9 mRNAs following transfection with ΨDNAs specific to these transcripts (ΨPPIA, ΨRPL4, and ΨPCSK9) in combination with either GFP (control) or AsCas12a (Fig. 6a, Supplementary Fig. S10 & Table 10). In the control cells transfected with GFP, we observed partial knockdown of the target transcripts, potentially due to low-level knockdown activity exerted by ΨDNAs alone. However, co-transfection of AsCas12a with the ΨDNAs led to a significant enhancement in transcript depletion. Specifically, ΨPPIA and ΨRPL4 resulted in an approximately 50% and 40% reduction in mRNA levels compared to the non-targeting (NT) control, respectively. Similarly, produced marked reductions in their respective transcript levels. These results demonstrate that while ΨDNA alone can induce partial knockdown, co-expression with AsCas12a substantially increases the efficiency and specificity of RNA transcript depletion, confirming the dependence of ΨDNA-mediated knockdown on the presence of AsCas12a.

**Fig. 6:**
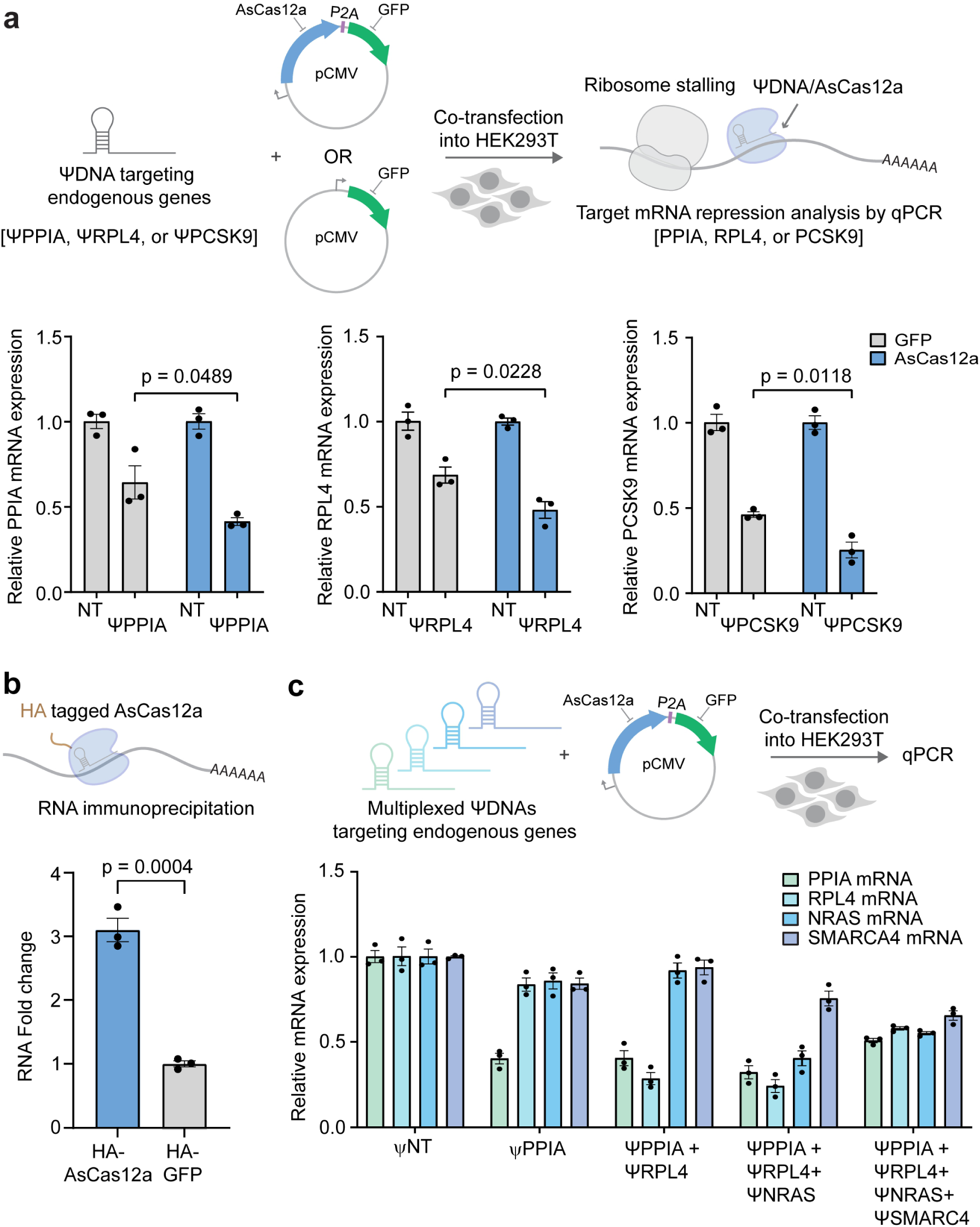
mRNA targeting of endogenous genes using ΨDNA-AsCas12a. **(a)** Relative mRNA expression levels of PPIA, RPL4, and PCSK9 transcripts in cells expressing either GFP (gray, control) or AsCas12a (green) with non-targeting ΨDNA (NT) or gene-specific ΨDNAs (ΨPPIA, ΨRPL4). Significant reduction in target gene expression is observed with ΨDNA-mediated knockdown when co-delivered with AsCas12a, demonstrating the specificity and efficiency of the RNA knockdown using ΨDNA-guided CRISPR-Cas12a system. Data are represented as mean ± S.E.M. **(b)** RNA Immunoprecipitation (RIP) shows significant enrichment of PPIA RNA in cells expressing HA-tagged AsCas12a compared to HA-GFP control. This demonstrates specific RNA targeting by the AsCas12a-ΨDNA complex. **(c)** Data shows multiplex knockdown of four endogenous transcripts PPIA, RPL4, NRAS, and SMARCA4 using a non-targeting ΨDNA (ΨNT) or various ΨDNA combinations as described: ΨPPIA (PPIA only), Ψ-Mix #1 (PPIA and RPL4), Ψ-Mix #2 (PPIA, RPL4 and NRAS) and Ψ-Mix #3 (PPIA, RPL4, NRAS and SMARCA4), showing differential and specific knockdown efficiency across gene targets. Results highlight the potential for multiplex RNA targeting using the ΨDNA-guided CRISPR-Cas12a platform. Data are presented as mean ± S.E.M. from *n* = 3 biological replicates.

To validate that AsCas12a is specifically recruiting and binding target RNA transcripts in the presence of ΨDNAs, we conducted RNA immunoprecipitation (RIP) using an HA-tagged version of AsCas12a. Cells delivered with either HA-GFP (control) or HA-AsCas12a and ΨPPIA were subjected to RIP, followed by qPCR analysis of the immunoprecipitated RNA to quantify the binding (Fig. 6b). We observed that PPIA transcripts were significantly enriched in samples expressing HA-AsCas12a compared to those expressing the control HA-GFP, demonstrating that AsCas12a binds directly to its targeted PPIA transcript when guided by ΨDNA. These results confirm that ΨDNA is facilitating direct interaction between AsCas12a and its endogenous RNA targets.

To evaluate the capability of ΨDNA-guided CRISPR-Cas12a for multiplex RNA transcript knockdown, we next co-delivered combinations of ΨDNAs targeting multiple endogenous transcripts, including PPIA, RPL4, NRAS, and SMARCA4. We co-transfected AsCas12a along with different ΨDNA combinations, including ΨPPIA (targeting PPIA only), Ψ-Mix #1 (targeting PPIA and RPL4), Ψ-Mix #2 (targeting PPIA, RPL4, and NRAS), and Ψ-Mix #3 (targeting PPIA, RPL4, NRAS, and SMARCA4) (Fig. 6c). We observed that individual ΨDNAs as well as multiplexed combinations (Ψ-Mix #1, #2, and #3) achieved efficient transcript depletion across all target RNAs. Specifically, Ψ-Mix #3 resulted in robust knockdown of PPIA, RPL4, NRAS, and SMARCA4 simultaneously, highlighting the potential of this system for targeting multiple transcripts in parallel. This multiplexed approach achieved a variable but notable reduction in the abundance of each transcript, with Ψ-Mix #3 showing the most substantial overall knockdown across all four targets.

Collectively, these findings demonstrate that the ΨDNA-guided CRISPR-Cas12a system enables precise and efficient depletion of endogenous RNA transcripts in a single or multiplexed manner. The system offers substantial versatility for RNA-targeting applications and holds potential for therapeutic interventions aimed at reducing the levels of disease-relevant RNA transcripts.

## Discussion

Overall, this study not only provides novel methodologies for RNA targeting but broadens the understanding of CRISPR-Cas systems, particularly Type V enzymes. Based on our binding affinity experiments, we showcased that ΨDNA can bind to Cas enzymes analogous to a crRNA. Additionally, the activation of trans-cleavage activity suggests that the Cas proteins not only bind to ΨDNA but also fold around the DNA guide similar to how they interact with crRNA. As the DNA-RNA heteroduplex binds to the Cas proteins, this interaction promotes the formation of the ternary complex and moves the RuvC domain to the final position to activate trans-cleavage activity. This mechanism is similar to how crRNA and dsDNA targets trigger non-specific ssDNA cleavage. ^3,10,14,32^. Moreover, the trans-cleavage activity of both enzymes, AsCas12a and Cas12i1, is greatly increased when the DNA guide has a 3’ handle that is situated in the same position as the 5’ scaffold of the crRNA. This evidence suggests the 3’ handle in ΨDNA further stabilizes the folding of the Cas protein and it is translated into a higher fluorescent signal. Additionally, we have observed no cis-cleavage or trans-cleavage of RNA, as Cas12 enzymes do not possess a HEPN domain capable of RNA degradation like Cas13 ^33^. Instead, the RuvC domain in Cas12 is specifically tailored to cleave DNA.

Current methods for RNA synthesis include solid-phase synthesis, *in vitro* transcription (IVT), and other enzymatic processes. Solid-phase synthesis of RNA is more expensive and time-consuming than that of DNA, requiring costly reagents, specialized chemistry, and extensive steps ^34^. IVT involves an initial DNA synthesis step, followed by enzymatic processes to produce RNA, adding to the overall cost and complexity. Additionally, RNA has several magnitude shorter shelf-life than DNA, necessitating low temperatures or specialized storage conditions to maintain stability. CRISPR-Cas nucleic acid detection platforms, such as SHERLOCK and DETECTR ^5,6^, predominantly rely on crRNA, making these technologies more expensive and less accessible. To address these challenges, this study aims to expand CRISPR-Cas research and develop more cost-efficient CRISPR diagnostics. We have established a DNA-guided RNA detection platform that effectively detects both short and long RNAs with high specificity and efficiency. In our work, we demonstrated the use of AsCas12a and ΨDNA in detecting RNA from HCV patient samples, achieving high efficiency and accuracy. This advancement not only makes CRISPR diagnostics more affordable but also enhances their accessibility, broadening their application in research and clinical diagnostics. Utilizing ΨDNA for RNA targeting not only reduces the expenses associated with RNA synthesis but also enables higher throughput for screening purposes. The rapid synthesis of ΨDNA libraries facilitates guide screening, which provides the flexibility to quickly generate and test various sequences for different targets. As we have demonstrated, large libraries of ΨDNA can be screened against numerous RNA targets (Fig. 3b, Supplementary Fig. S1) and various scaffold sequences can be tested to rapidly optimize the construct (Fig. 3e). This efficiency in time and cost is particularly advantageous in high-throughput applications to allow faster iteration of guides and optimize experimental conditions. Thus, the use of ΨDNA represents a practical and economical alternative for large-scale studies involving RNA detection.

Lastly, as the CRISPR-Cas toolkit continues to evolve, we introduce a new platform for RNA targeting with a novel mechanism for Cas12 enzymes. CRISPR-Cas systems, widely regarded as powerful tools for DNA and RNA editing, have been extensively engineered for genome editing within cells. Our study demonstrates that AsCas12a can complex with ΨDNA and target mRNA inside cells, leading to reduced mRNA levels and lower protein production through translational repression. Importantly, since this DNA-guided system lacks RNA cis- or trans-cleavage activity, we have shown that the ΨDNA-Cas12 complex is effective for transient gene silencing and holds potential for RNA editing within cellular environments as other researchers have achieved with inactive variants of Cas13 ^35–37^. Furthermore, we have demonstrated multiplexed gene silencing through four different targets which holds great potential for multiplexed gene screening similar to previous work with Cas13 ^38^. In the multiplex knockdown system, the knockdown efficiency observed for the Ψ-Mix #3 (4 targets) group was lower than that of the Ψ-Mix #2 (3 targets) group. This discrepancy is attributed to the transfection of an equivalent total amount of Ψ-DNA, leading to a reduced allocation of Ψ-DNA per target in 4 targets condition. Enhancing this system could be achieved by increasing the overall Ψ-DNA transfection amount, thereby facilitating more potential for knockdown of more target genes. This innovation not only extends the functionality of Cas12 beyond conventional applications but also boosts the versatility and efficiency of CRISPR-based technologies in both research and therapeutic settings. Furthermore, the Cas12-ΨDNA construct holds potential for future studies as it can be fused to other functional proteins, offering additional modularity for tailored applications in RNA editing and manipulation, such as localized RNA labeling or base modification within cells.

### Limitations of the study

Despite the advantages provided by the programmable DNA-guided RNA targeting platform, a key limitation holding back its full potential is the low detection sensitivity without pre-amplification. While we demonstrated picomolar level detection with our assay, this is not enough to accurately detect a wide range of patient diseases that often need the sub-femtomolar to attomolar level of detection. Even though pre-amplification techniques such as RT-PCR or RT-RPA improve the detection limit, they add significantly to the cost and complexity of the detection process and are not ideal. Additionally, further optimization of gene repression within cells could enhance the system’s effectiveness. As observed *in vitro*, the construct’s flexibility allows for the incorporation of various scaffold sequences, which could improve its functionality and increase activity in a cellular environment. Furthermore, fusing different domains to Cas12 proteins can further improve and expand the RNA editing capabilities.

## Acknowledgements

We extend our gratitude to the HCV-TARGET consortium, particularly Dr. David Nelson and Lauren Morelli, for their expert guidance and provision of clinical samples. We also thank the members of the Jain and Wang labs for valuable discussions and the University of Florida (UF) Health Cancer Center for their support. Finally, we thank Dr. Carl Denard and his lab members in the Department of Chemical Engineering and UF ICBR Monoclonal Antibody Core (RRID:SCR_019147) for their guidance on BLI experiments and UF ICBR Cytometry Core (RRID:SCR_019119) for their help with flow cytometry experiments. This work was funded by the University of Florida, the UF Herbert Wertheim College of Engineering, the Shah Foundation Endowment Funds, NIH-NIAID (R21AI156321, R21AI168795, and R61AI181016), and NIH-NIGMS (R35GM147788). The funding sources had no role in study design, data collection, analysis, interpretation, or manuscript preparation.

## Data Availability

All data supporting the findings of this study are provided within the Article and Supplementary Files. Additional data may be obtained from the corresponding author, P.K.J., upon reasonable request.

