## Supplementary File for "DNA-guided CRISPR/Cas12 for RNA targeting"

**The PDF file includes:**

Materials and Methods
Figs. S1 to S11
Tables S1 to S10
References

### Materials and Methods

Plasmid Construction

Plasmids encoding Lb, As, Er and other Cas12a variants and the BrCas12b enzyme were built following the protocol outlined in our earlier publications [22,23,28,39-41]. Plasmid containing E. coli codon-optimized Cas12i1 and Cas12i2 gene was obtained from Addgene, a gift from ArborBiotechnologies (Plasmid #120882 & #120883).

For mammalian cell transfection, NLS was removed from the AsCas12a-GFP plasmid (Addgene #160140) through a KLD reaction. mCherry plasmid was constructed by inserting the mCherry gene into a pCMV vector with no CMV enhancer sequence.

Protein Expression and Purification

Rosetta bacterial colonies harboring the plasmid for protein production were cultured at 37°C overnight on agar. Select colonies were transferred from the agar to 10 mL LB medium (Fisher Scientific, Catalog #BP9723-500) for 12-hour incubation. The culture was then expanded to 1.5 L TB medium (MP Bio #113046042), growing until an optical density (OD) of 0.6-0.8 was reached. The culture was cooled for 45-60 minutes before adding Isopropyl β-d-1-thiogalactopyranoside (IPTG) to a concentration of 0.5 mM, and growth continued for 14-18 hours at 16°C.

Cell harvesting involved centrifugation at 10,000 x g for 5 minutes. The pellet was reconstituted in Lysis Buffer (500 mM NaCl, 50 mM Tris-HCl, pH 7.5, 20 mM Imidazole, 0.5 mM TCEP, 1 mM PMSF, 0.25 mg/mL Lysozyme, DNase I). This suspension underwent sonication and further centrifugation at 39,800 xg for 30 minutes. The resulting lysate was passed through a 0.22 µm syringe filter (Cytiva, Catalog #9913-2504) and introduced into a 5 ml Histrap FF column (Cytiva, Catalog #17525501, with Ni2+ replaced by Co2+) linked to a BioLogic DuoFlow™ FPLC system (Bio-rad).

Proteins were eluted using Buffer B (500 mM NaCl, 50 mM Tris-HCl, pH 7.5, 250 mM Imidazole, 0.5 mM TCEP). For all proteins except Cas12i1 and Cas12i2, the eluted fractions were merged and dialyzed in a 10 kDa – 14 kDa MWCO bag with TEV protease (sourced from David Waugh, Addgene #8827), and prepared internally. The bag was immersed in Dialysis Buffer (500 mM NaCl, 50 mM HEPES, pH 7, 5 mM MgCl2, 2 mM DTT) and stirred gently at 4°C overnight.

The protein blend was concentrated to approximately 10 mL using a 30 kDa MWCO Vivaspin® 20 concentrator. This concentrate was then balanced with 10 mL of Buffer C (150 mM NaCl, 50 mM HEPES, pH 7, 0.5 mM TCEP). It was processed through a 1 mL Hitrap Heparin HP column (prepped with Buffer C) using the BioLogic DuoFlow™ FPLC system (Bio-rad). A gradient flow alternating between Buffer C and Elution Buffer D (2000 mM NaCl, 50 mM HEPES, pH 7, 0.5 mM TCEP) facilitated protein elution. Further size-exclusion chromatography was employed as necessary. The protein passed through a HiLoad® 16/600 Superdex® column (Cytiva, Catalog #28989335), and the highest purity fractions were pooled, concentrated with a 30 kDa MWCO Vivaspin® 20, and flash-frozen in liquid nitrogen for storage at –80°C.

Oligonucleotide preparation

All single-stranded DNA and RNA oligos including guide RNA, guide DNA, target activators, primers, and fluorescent reporters were obtained from Integrated DNA Technologies (IDT) and diluted in 1x TE Buffer (10 mM Tris, 0.1 mM EDTA, pH 7.5). For generating long RNA target mimics of HIV, Zika, Dengue and HCV RNA, dsDNA gene fragments containing a T7 promoter region were ordered from Twist Biosciences and in vitro transcribed using HiScribe® T7 High Yield RNA Synthesis Kit (NEB# E2040S) to generate the long RNA fragments.

CRISPR-Cas based fluorescence detection assay

All detection assays using fluorescence were performed in black 384-well plates with a low-volume, flat-bottom design. The crRNA-Cas12 complexes were prepared by mixing in NEB 2.1 buffer and nuclease free water, followed by a 10-minute room temperature incubation. These crRNA-Cas12 preparations were then combined with 250-500 nM of a fluorescent quenched (FQ) reporter and an appropriate amount of the target activator to achieve a reaction volume of 40 µL. The 384-well plate was subsequently placed in a BioTek Synergy fluorescence microplate reader, incubated at temperature of 37 °C for 60 minutes. Fluorescence intensity readings for a FAM-labeled reporter were recorded at 483/20 nm and 530/20 nm excitation/emission wavelengths at 2.5-minute intervals. Standard concentrations in these assays, unless otherwise noted, were 50 nM Cas enzyme (AsCas12a/Cas12i), 100 nM crRNA or ΨDNA, and 25 nM target activator, unless otherwise specified.

Electrophoretic Mobility Shift Assay (EMSA)

EMSA gel was performed by mixing 100 nM of Cas12 protein (AsCas12a or Cas12i1), 100 nM of guide (crRNA or ΨDNA) and 100 nM target (ssDNA or ssRNA) in NEB 2.1 buffer, and incubating the reaction at 4 °C for 30 min. Then, 1 µL of 5x TBE Hi-Density Sample Buffer (Invitrogen #LC6678) was added to 9 µL of sample. The samples were loaded in a native PAGE DNA Retardation Gel (Invitrogen #EC6365BOX) and ran at 200V for 30 min. Later PAGE gel was stained with SYBR Gold (Invitrogen # S11494) and imaged on Amersham Typhoon.

Bio-layer Interferometry

The biolayer interferometry (BLI) analysis of the binding interaction between Cas enzymes and DNA or RNA based guides was performed using the GatorBio instrument, employing biotin-coated guides and streptavidin probes from the Flex SA Kit (GatorBio #350001). Priming reagent was switched to NEB 2.1 buffer with 0.05% Triton X. DNA and RNA oligos were synthesized with a 3' biotin modification through Integrated DNA Technologies (IDT).

The BLI measurements were conducted on the GatorBio system, which allows real-time, label-free analysis of biomolecular interactions. The assay setup involved two key phases: association and dissociation. During the association phase, the streptavidin probes loaded with biotinylated guides at 50 nM were transferred to wells containing the Cas enzyme (Cas12i1 or AsCas12a) at seven concentrations (1 nM, 2.5 nM, 5 nM, 10 nM,25 nM, 50 nM, and 100 nM). The binding of the Cas enzyme to the immobilized guide was monitored in real-time over a 10-minute period. In the subsequent dissociation phase, the sensors were moved to wells containing only the assay buffer to observe the dissociation of the Cas enzyme from the immobilized guide over a 10-minute period.

The BLI data was processed and analyzed using GatorBio software. Binding curves were generated for each concentration of the Cas enzyme, and the association ($k_{on}$) and dissociation ($k_{off}$) rate constants were determined by fitting the data to a 1:1 binding model. The equilibrium dissociation constant ($K_d$) was calculated using a Michaels-Menten regression curve to fit the response curve at different concentrations. Comparative analyses between DNA and RNA guides were conducted to evaluate the specificity and affinity of the Cas enzyme for each type of guide. Sensors without immobilized guides served as negative controls to confirm the specificity of the streptavidin-biotin interaction and the subsequent guide-Cas binding.

In vitro transcription

*In vitro* transcription of long HIV and HCV RNA fragments was done using HiScribe® T7 High Yield RNA Synthesis Kit (NEB# E2040S) following manufacturer’s protocol and purified using Monarch RNA cleanup kit (NEB# T2030L).

Endogenous mRNA detection

Total RNA of 1 x $10^6$ HEK293T cells (ATCC #CRL-3216) was extracted using Monarch Total RNA Miniprep Kit (NEB #T2010S). Then, cDNA was synthesized using PhotoScript II First Strand cDNA Synthesis Kit (NEB #E6560) with Oligo-dT primers. Lastly, 17 endogenous genes were amplified using KOD One PCR Master Mix (Toyobo #KMM-201) and *in vitro* transcribed as described above. Fir detection, 2 µL of product were added to a CRISPR-Cas based fluorescence detection assay.

Patient sample collection

The collection and processing of patient samples were approved by the University of Florida Institutional Review Board (IRB202200294). For clinical validation, a total of 20 human serum samples were obtained from patients with Hepatitis C collected under the HCV-TARGET program, which also supplied the UI/ml of each sample reported in Fig. 4. Healthy serum samples were obtained from Boca Biolistics.

Patient sample extraction

The extraction of viral RNA from serum samples was carried out using the Quick-DNA/RNA Viral MagBead Kit (Zymo #R2140). Briefly, 10 µL of Proteinase K (20 mg/mL) was added to 200 µL of the serum samples in 1.5 mL centrifuge tubes, followed by incubation at room temperature for 15 minutes. Afterward, DNA/RNA Shield™ (2x concentrate) was mixed with the serum sample containing Proteinase K at a 1:1 ratio. The resulting mixture was then supplemented with 800 µL of Viral DNA/RNA Buffer, followed by the addition of 20 µL of Magbinding Beads™. The mixture was vortexed for 10 minutes, and then the tubes were placed on a magnetic stand to separate the beads. The supernatant was carefully removed, and the beads were sequentially washed with 250 µL of MagBead DNA/RNA Wash 1, 250 µL of MagBead DNA/RNA Wash 2, and two rounds of 250 µL of 100% ethanol. After allowing the beads to air-dry for 10 minutes, the DNA/RNA was eluted with 30-60 µL of DNase/RNase-Free water and subsequently used for downstream analysis.

Patient sample validation

After extraction, samples were amplified by adding 1 µL of extracted viral RNA to a 50 µL SuperScript™ IV One-Step RT PCR reaction (Invitrogen #12594025) and performed thermocycling according to manufacturer's instructions. After amplification, 1 µL of product was added to a T7 and AsCas12a reaction master mix for RNA detection. This master mix contained 1x NEB 2.1 buffer, 62.5 nM AsCas12a, 112.5 nM ΨDNA (IDT), 500 nM FQ reporter (IDT), 1mM rNTPs Mix (NEB #N0466), 2 U/ml of RNase Inhibitor (NEB #M0314), and 12.5 U/ml of NxGen T7 RNA Polymerase (Biosearch #30221-1) in 20 µL reactions. Samples were then loaded into a 384-well plate which was subsequently placed in a BioTek Synergy fluorescence microplate reader, incubated at temperature of 37 °C for 60 minutes. Fluorescence intensity readings for a FAM-labeled reporter were recorded at 483/20 nm and 530/20 nm excitation/emission wavelengths at 2.5-minute intervals.

Illumina NGS sequencing for patient samples

Extracted samples were amplified through a SuperScript™ IV One-Step RT PCR reaction as the step above. A second round of PCR was perfomed using Q5 DNA Polymerase to append Illumina barcodes. Samples were then pooled together, gel extracted and loaded into a Illumina MiSeqDx with a MiSeq Reagent Nano Kit v2 (Illumina #MS-101-1001). Bowtie2, Samtools, and JBrowse were used to align the sequencing output to an HCV reference genome.

Mammalian cell culture

HEK293T cells (ATCC #CRL-3216) were cultured in DMEM high glucose GlutaMAX™ supplement pyruvate (Gibco #10569010), 10% Fetal Bovine Serum (Gibco #A3160902) and 1X Penicillin-Streptomycin. Cells were incubated at 37°C and 5% $CO_2$.

Mammalian cell transfection

For RNA extraction and qPCR, plasmids were co-transfected with different ΨDNAs into HEK293T cells using TransIT-X2® transfection reagent (Mirus Bio #MIR6000). $5x10^5$ cells per well were seeded in 48-well plates 48 hours before transfection.

For mCherry reporter system, 200 ng of mCherry plasmid, 350 ng of AsCas12a-GFP (Addgene #160140) or GFP only (Addgene #133962) plasmid, 400 ng of ΨDNA and 2 µL of TransIT-X2® were added to 50 µL of Opti-MEM™ Reduced Serum Medium (Gibco, Cat# 31985062). Complexes were allowed to form for 25 minutes and posteriorly added dropwise to each well. For the control reaction with no ΨDNA, to normalize transfection efficiency among all samples 400 ng of a short-randomized DNA were added to the reaction instead of ΨDNA. Transfected cells were harvested 16 hours post-transfection for flow cytometry and gene expression analysis.

For endogenous gene knockdown, 600 ng of AsCas12a-GFP or GFP-only plasmid, 400 ng of total ΨDNA, and 2 µL of TransIT-X2® were added to 50 µL of Opti-MEM™ Reduced Serum Medium. For multiplex gene targeting, different ΨDNAs were equally mixed before being transfected. Cells were harvested 16 hours post-transfection for gene expression analysis.

For RIP-qPCR, $2x10^6$ per well HEK293T cells were plated in 48-well plates 24 hours before transfection. 1.33 µg of AsCas12a-GFP or HA-GFP, 1.07 µg ΨDNA, and 6 µL of TransIT-X2® were added to 250 µL of Opti-MEM™ Reduced Serum Medium. Transfected cells were harvested 16 hours post-transfection.

Flow cytometry for quantification of mCherry expression

16 hours after cells were transfected as described above, these were trypsinized with 1X Trypsin-EDTA (Gibco #15400054). Then the cells were resuspended in FluoroBrite™ DMEM with 10% FBS. Each sample was passed through a 35 um cell strainer (FALCON #352235). Cells were then analyzed in a Beckman Coulter CytoFLEX LX flow cytometer. FCS files were then analyzed in FloJo™ v.10.10 to obtain the MFI of mCherry expression in GFP-positive cells. Before analyzing samples compensation for GFP and mCherry was performed using GFP cells, mCherry cells, and GFP & mCherry BrightComp eBeads™ (Invitrogen #A10514 and #A54743). Gating illustrated in Fig. S12.

RT-qPCR for relative quantification of mCherry mRNA

Total RNA of samples was extracted using Monarch Total RNA Miniprep Kit (NEB #T2010S) as per manufacturer's instructions. Extracted RNA was later added to the RT-qPCR mix Luna® Universal Probe One-Step RT-qPCR Kit (NEB #E3006). The reaction was performed on a Applied Biosciences QuantStudio™ 5 Real-Time PCR System and multiplexed with FAM probe for mCherry and Cy5 probe for GAPDH as the endogenous control gene. mCherry primers and probes were designed using PrimeQuest™ Tool from IDT and GAPDH primers and probes are from Asahi-Ozaki *et al* [42]. For others genes, primers and probes were ordered from Thermo Fisher. Fold-change was calculated relative to non-target ΨDNA using the ddCt method. One-way or two-way ANOVA with multiple comparison correction was performed to assess the statistical significance of transcript changes, using Prism 10.

RNA immunoprecipitation and Quantitative PCR (RIP-qPCR)

RNA immunoprecipitation was performed as previously described [43]. HA-AsCas12a or HA-GFP and Ψ-PPIA transfected HEK293T cells were fixed with 1% paraformaldehyde (ChemCruz # sc281692) for 15 minutes at room temperature. After fixation, the paraformaldehyde was removed, and 125 mM glycine in PBS was added to quench the crosslinking, followed by a 10-minute incubation. Cells were washed twice with ice-cold PBS, harvested by scraping, and the cell suspension was centrifuged at 1000 g for 5 minutes to pellet the cells. The cell pellets were re-suspended in 2 volumes of lysis buffer (150 mM NaCl, 10 mM Tris-HCl pH 7.6, 2 mM EDTA, 0.5% NP-40, 0.5 mM DTT, 400 U/ml RNase inhibitor (NEB #M0314S)) and mixed thoroughly. The lysates were incubated on ice for 20 minutes. Insoluble material was pelleted by centrifugation at 15,000 g for 15 minutes at 4°C, and the supernatant, containing the cleared lysate, was used for pulldown with Protein G Agarose beads (Sigma, 16-201). Set aside 30 µl of the supernatant as input.

To conjugate antibodies to agarose beads, 20 µL of Protein G Agarose beads per sample for immunoprecipitation were washed twice and resuspended in 300 µL of wash buffer (150 mM NaCl, 50 mM Tris-HCl, pH 7.6, 2 mM EDTA, 0.05% NP-40, 0.5 mM DTT, 200 U/mL RNase inhibitor). Then, 2 µL of anti-HA antibody (Cell Signaling Technology #3724) was added. The sample was incubated for 2 hours at 4°C on a rotator to allow the antibody to conjugate to the beads. After incubation, the beads were pelleted by centrifugation, the supernatant was removed, and 100 µL of sample lysate was added to the beads and rotated overnight at 4°C.

After incubation with sample lysate, beads were pelleted, washed three times with High-salt buffer (300 mM NaCl, 50 mM Tris-HCl pH 7.6, 2 mM EDTA, 0.05% NP-40, 0.5 mM DTT, 200 U ml–1 RNase inhibitor), and then resuspend with SDS solution (1% SDS, 10 mM EDTA (pH 8.0), 50 mM Tris-HCl, pH 7.4, 200 U/ml RNase inhibitor). Proteins were then digested by addition of Proteinase K (NEB #P8102S) to a final concentration of 1.2 mg/ml and incubated at 55°C for 4 hours. TRIzol (Invitrogen # 15596018) was added for denaturation and RNA purification. Purified

RNA was reverse transcribed and amplified by Luna® Universal Probe One-Step RT-qPCR Kit. Fold change was calculated relative to input using the ddCt method.

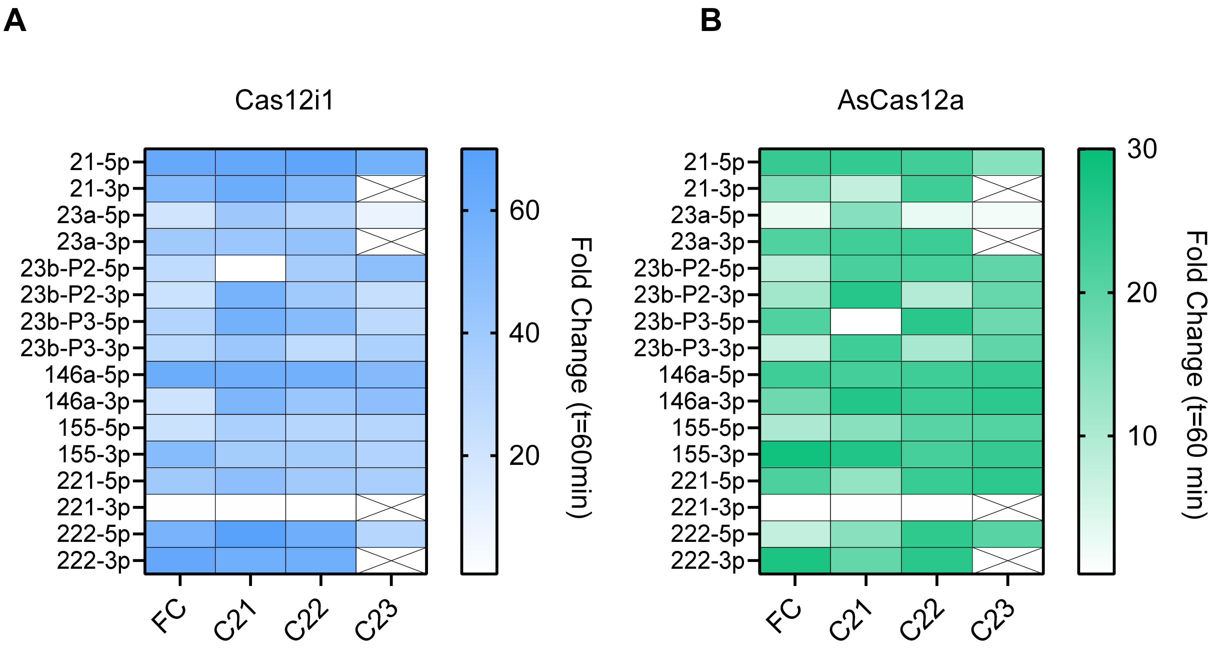


**Fig. S1. Detection of 16 miRNA targets with Cas12 and ΨDNA.** (**A**) Heatmap of miRNA detection with ΨDNA and Cas12i1 and (**B**) AsCas12a. (**A, B**) FC denotes that the spacer region of ΨDNA is fully complementary to the miRNA; C21 denotes the spacer region of ΨDNA targets 1-21 nt of miRNA from 5' end; C22 denotes the spacer region of ΨDNA targets 2-22 nt of miRNA; C23 represents the spacer region of ΨDNA target 3-23 nt of miRNA if miRNA is 23 bases or longer.

**A**

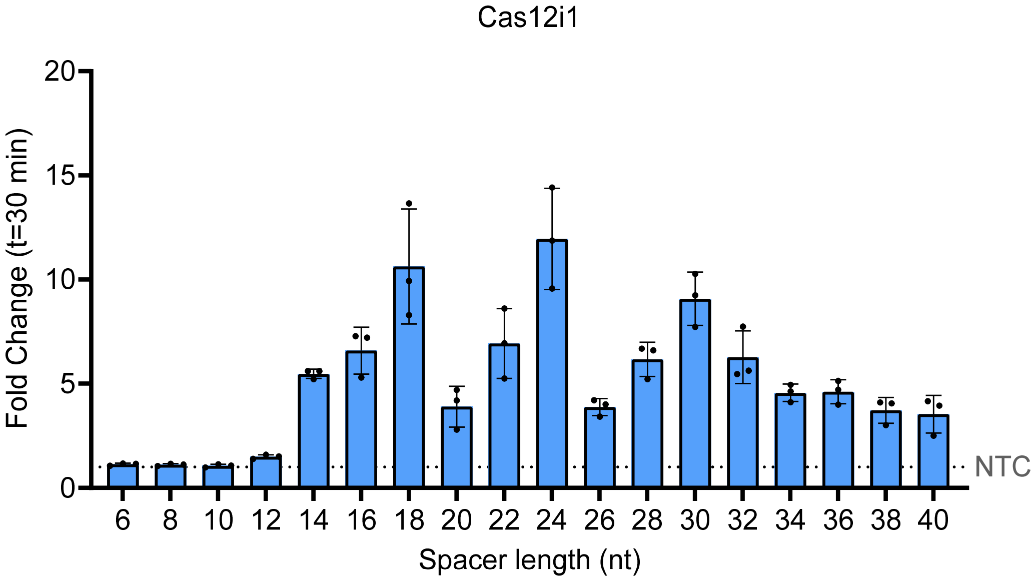


**B**

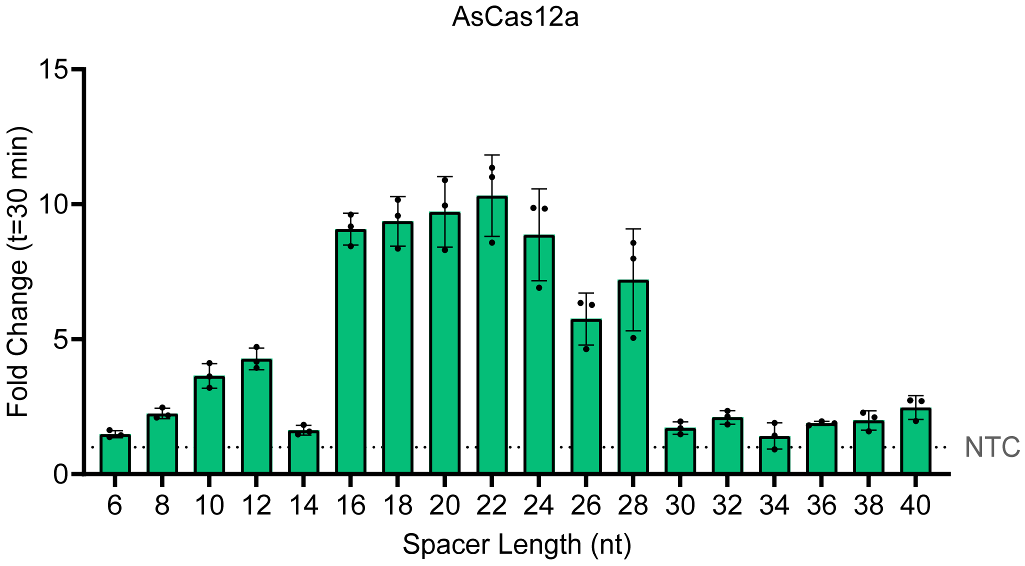


**Fig. S2. Trans-cleavage activity of ΨDNA with AsCas12a and Cas12i1 with varying spacer length.** (**A**) Trans-cleavage activity of Cas12i1 (blue) with ΨDNAs of different spacer lengths. (**B**) Trans-cleavage activity of AsCas12a (green) with ΨDNAs of different spacer lengths. (**A, B**) Fluorescence is measured at 30 min. Fold change of fluorescence intensity normalized to No Target Control (NTC). (n=3 technical replicates; bars represent mean ± S.D.). Target is 50 nucleotides long.

**A**

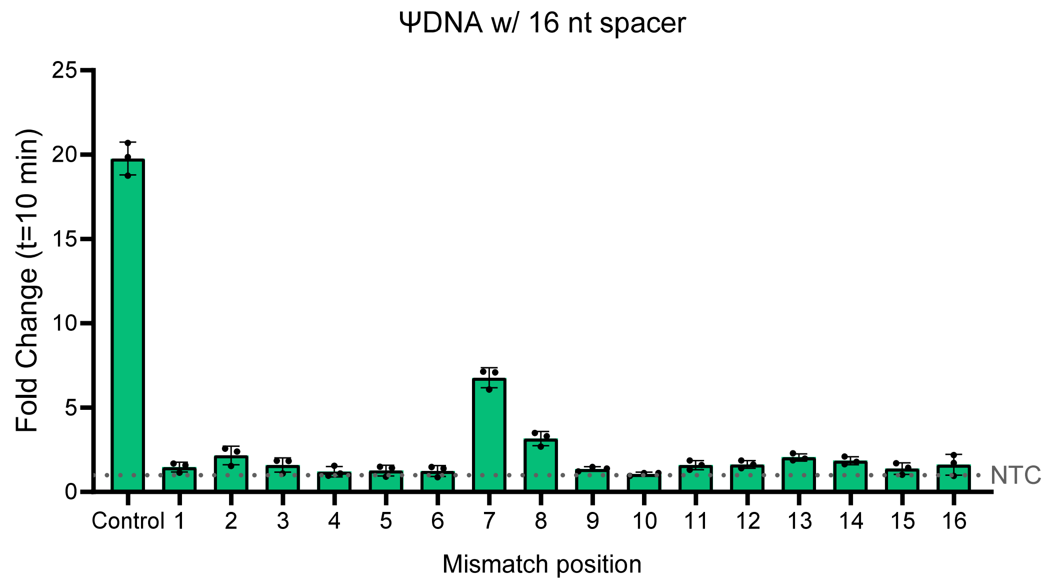


**B**

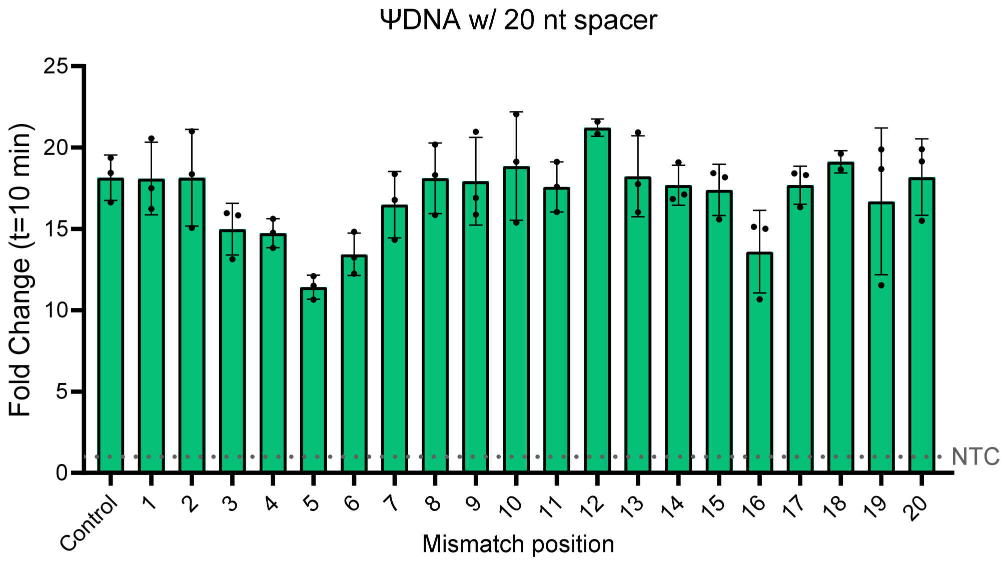


**Fig S3. Trans-cleavage activity of AsCas12a with ΨDNA with mismatches at different positions.** (**A**) Trans-cleavage activity of AsCas12a with 16-mer spacer ΨDNA with mismatches at different positions. (**B**) Trans-cleavage activity of AsCas12a with 20-mer spacer ΨDNA with mismatches at different positions. (**A, B**) Fluorescence is measured at 10 min. Fold change of fluorescence intensity normalized to No Target Control (NTC). (n=3 technical replicates; bars represent mean ± S.D.). RNA target matches spacer length.

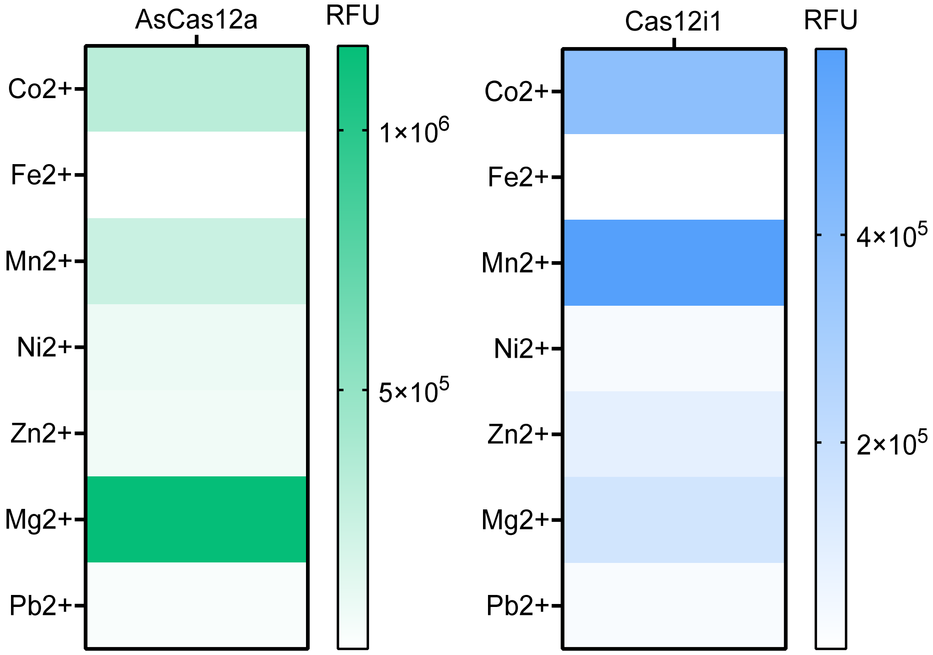


**Fig. S4. Effect of different divalent cations on ΨDNA induced trans-cleavage for RNA detection.** $Mg^{2+}$ in NEB2.1 buffer was replaced by different divalent cations. Heatmaps demonstrate AsCas12a has the highest activity with $Mg^{2+}$ and Cas12i1 has improved activity with $Co^{2+}$ and $Mn^{2+}$.

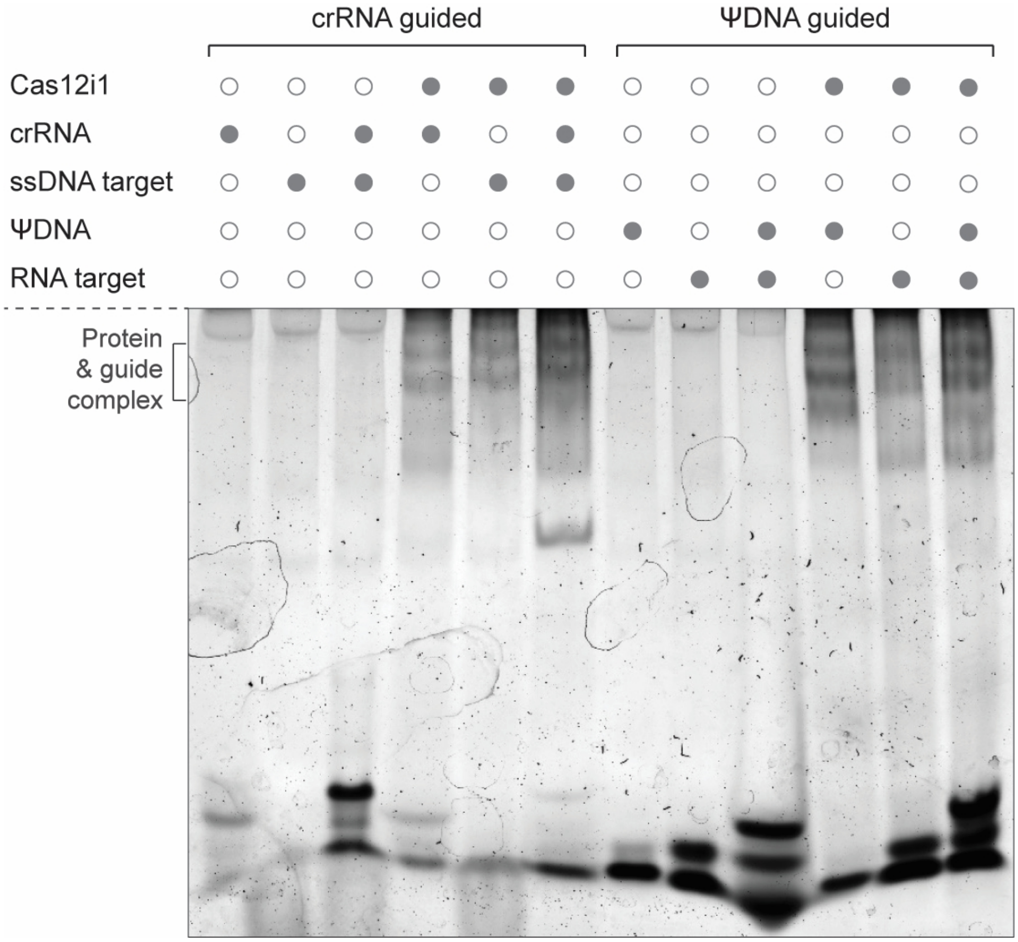


**Fig. S5. Electrophoretic Mobility Shift Assay (EMSA) for Cas12i1 and ΨDNA.** EMSA comparing the complex formation of Cas12i1 with either a canonical crRNA guide targeting a ssDNA target (lanes 1-6) versus a ΨDNA guide targeting an RNA target (lanes 7-12). This gel demonstrates the ability of ΨDNA guides to form stable complexes with Cas12i1 and target RNA.

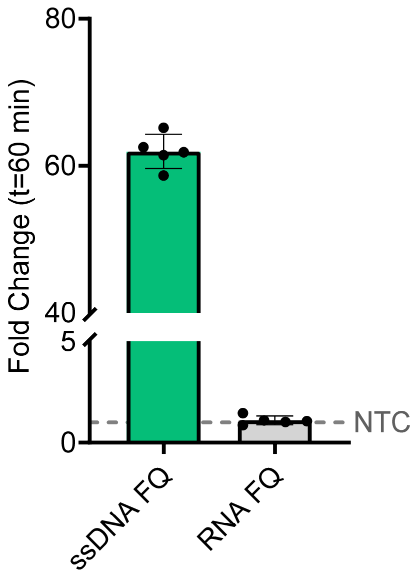


**Fig. S6. AsCas12a trans-cleavage activity with ΨDNA against ssDNA and RNA reporters.** ΨDNA induced trans-cleavage for RNA detection. Non-specific collateral cleavage of nucleic acids tested with ssDNA and RNA reporters. Fold change of fluorescence intensity normalized to No Target Control (NTC). (*n* = 5, technical replicates; bars represent mean ± S.D.)

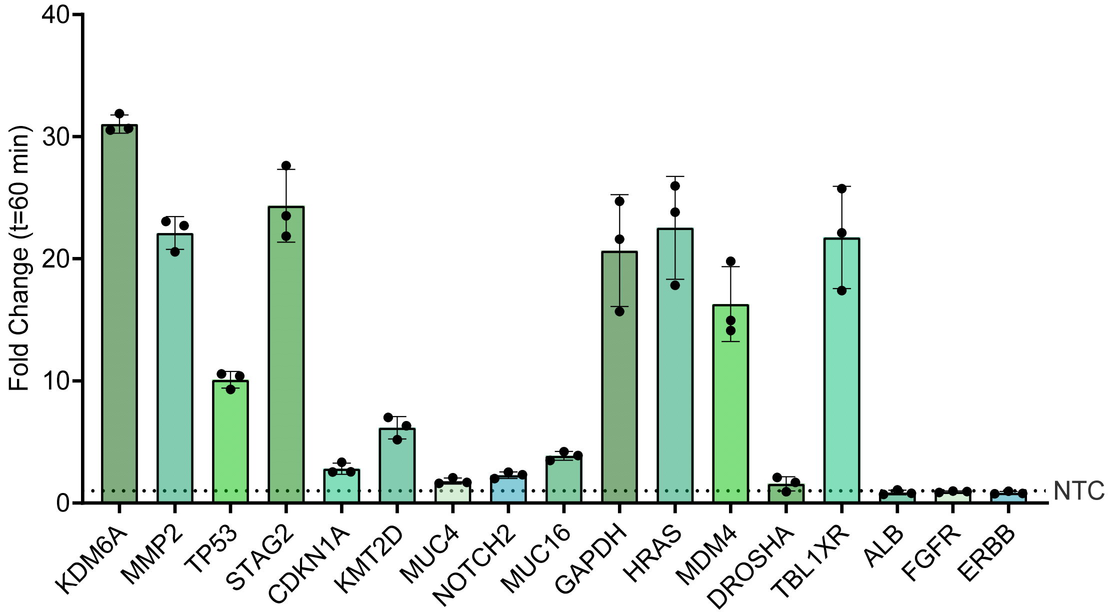


**Fig. S7. Endogenous mRNAs detection by AsCas12a and ΨDNA.** Fluorescence measurements for mRNA detection with ΨDNA and AsCas12a. HEK293T endogenous mRNAs were amplified by RT-PCR and *in vitro* transcribed. A total of 17 mRNA targets were tested. Fold change of fluorescence intensity normalized to No Target Control (NTC). (n=3 technical replicates; bars represent mean ± S.D.).

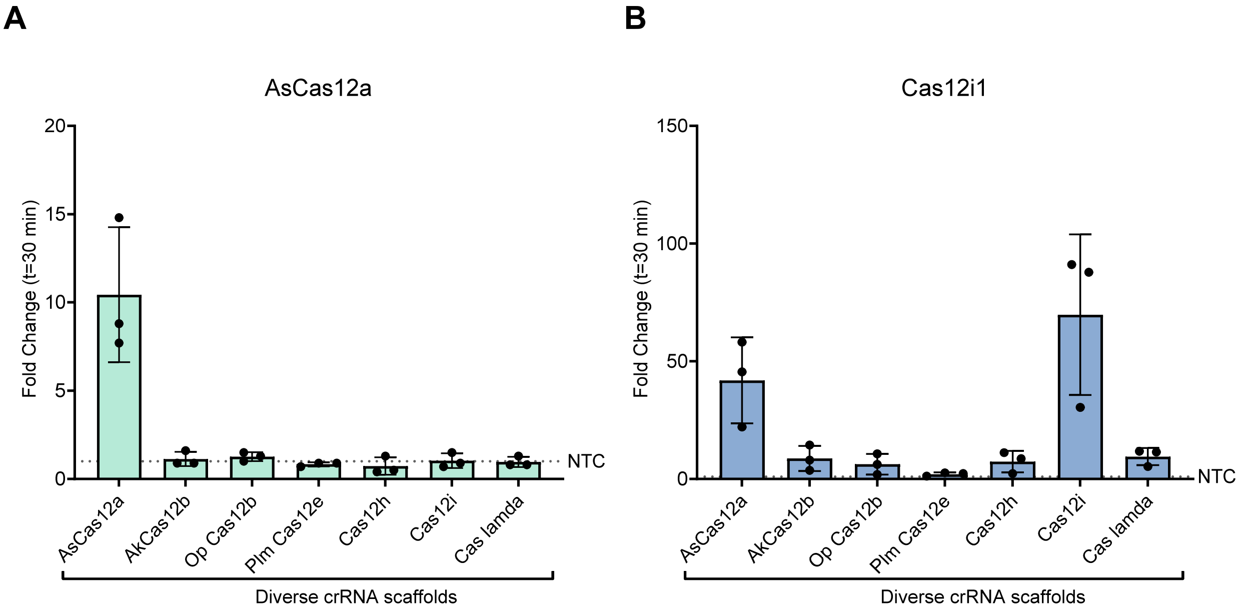


**Fig. S8. Trans-cleavage by Cas12 and crRNA with different scaffolds.** Cas12i1 (**A**) and AsCas12a (**B**) trans-cleavage activity with crRNAs of different scaffolds to detect a ssDNA target. The scaffolds chosen were the sequences that showed high trans-cleavage activity when used as the 3' handle of ΨDNA to detect RNA. Fold change of fluorescence intensity normalized to No Target Control (NTC). (n=3 technical replicates; bars represent mean ± S.D.)

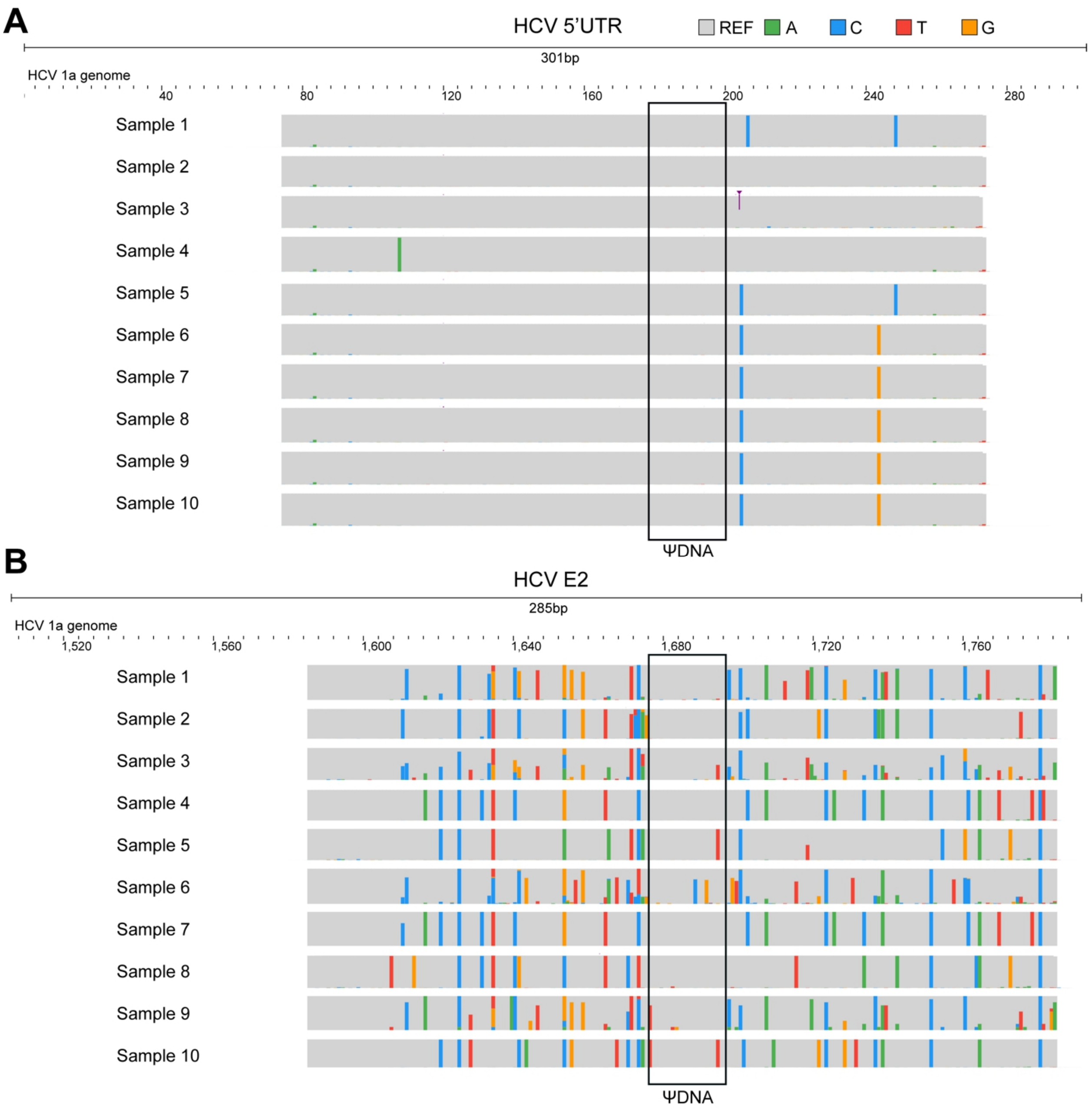


**Fig. S9. Sequence alignments of HCV patient samples by NGS**. Sequence alignments of (**A**) 5'UTR and E2 (**B**) regions of HCV patient samples. (**A, B**) Gray sections represent matched bases to the reference genome and colored sections represent mismatched bases. Alignments were visualized by JBrowse.

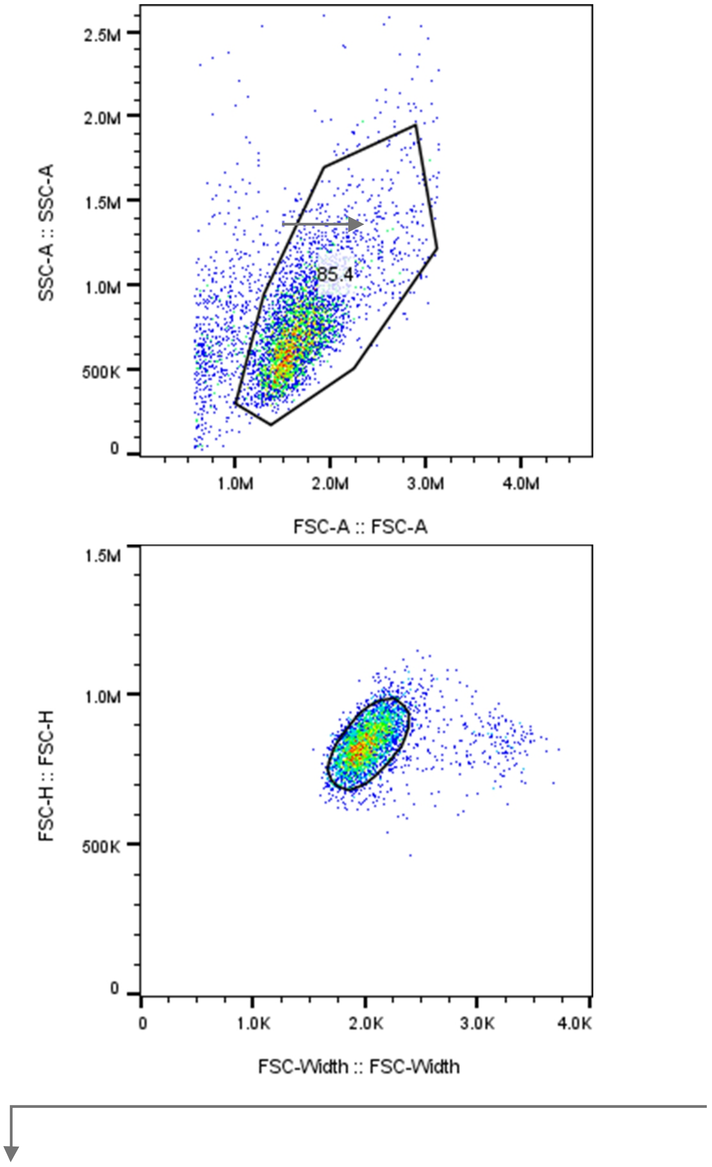
2.5M
2.0M
1.5M
1.0M
500K
0
SSC-A :: SSC-A
85.4
1.0M
2.0M
3.0M
4.0M
FSC-A :: FSC-A
1.5M
1.0M
500K
0
FSC-H :: FSC-H
0
1.0K
2.0K
3.0K
4.0K
FSC-Width :: FSC-Width

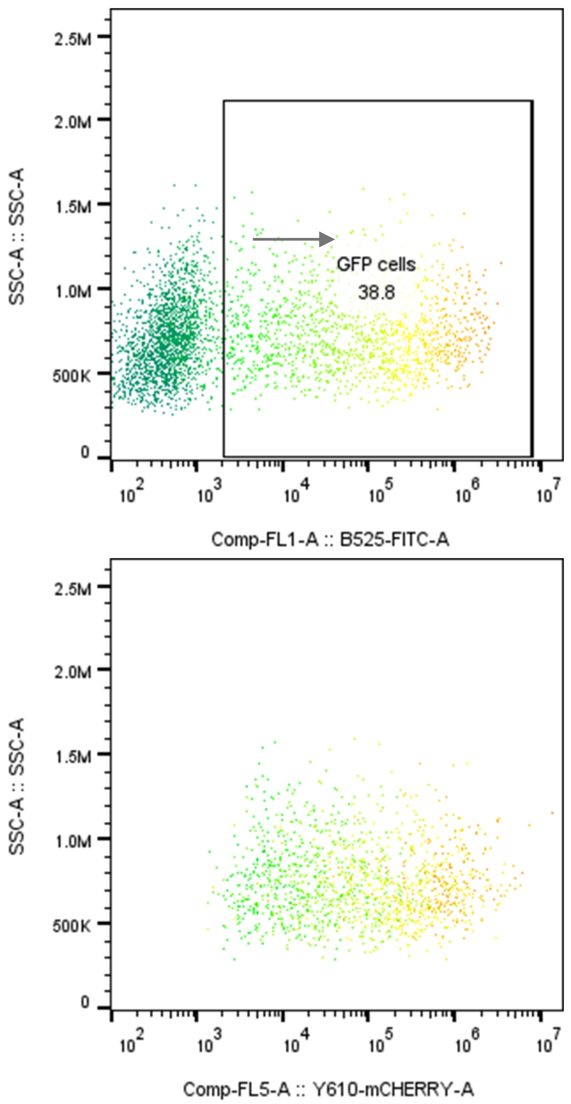


**Fig. S10. FACS gating strategy for mCherry gene silencing experiments**. Gating to determine the MFI of mCherry and GFP for all samples in Fig. 5. First gates exclude cell debris and only select for singlets. Later GFP-positive cells are gated and the MFI of GFP is calculated. Finally, all GFP-positive cells are plotted to find the MFI of mCherry.

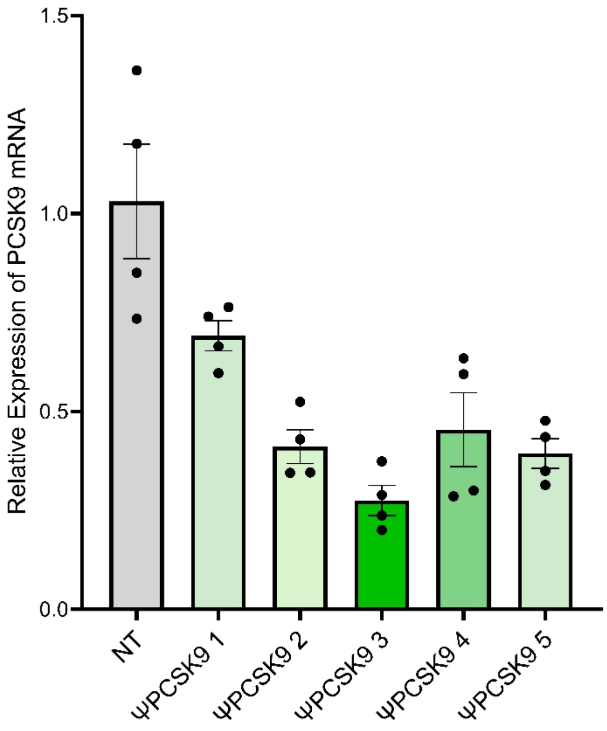


**Fig. S11. Screen of PCSK9 ΨDNA for reduction of mRNA.** Relative mRNA expression levels of PCSK9 transcripts in cells for 5 different ΨDNAs compared to an NT guide. Significant reduction in target gene expression is observed with ΨDNA-mediated knockdown when co-delivered with AsCas12a,

**Table S1.** Protein amino acid sequences

| Protein | Amino Acid Sequence |
|---|---|
| AsCas12a | MTQFEGFTNLYQVSKTLRFELIPQGKTLKHIQEQGFIEEDKARNDHYKELKPIIDRIYKTYAD<br>QCLQLVQLDWENLSAAIDSYRKEKTEETRNALIEEQATYRNAIHDYFIGRTDNLTDAINKR<br>HAEIYKGLFKAELFNGKVLKQLGTVTTTEHENALLRSFDKFTTYFSGFYENRKNVFSAEDIS<br>TAIPHRIVQDNFPKFKENCHIFTRLITAVPSLREHFENVKKAIGIFVSTSIEEVFSFPFYNQLLT<br>QTQIDLYNQLLGGISREAGTEKIKGLNEVLNLAIQKNDETAHIIASLPHRFIPLFKQILSDRNT<br>LSFILEEFKSDEEVIQSFCKYKTLLRNENVLETAEALFNELNSIDLTHIFISHKKLETISSALCD<br>HWDTLRNALYERRISELTGKITKSAKEKVQRSLKHEDINLQEIISAAGKELSEAFKQKTSEIL<br>SHAHAALDQPLPTTLKKQEEKEILKSQLDSLLGLYHLLDWFAVDESNEVDPEFSARLTGIKL<br>EMEPSLSFYNKARNYATKKPYSVEKFKLNFQMPTLASGWDVNKEKNNGAILFVKNGLYYL<br>GIMPKQKGRYKALSFEPTEKTSEGFDKMYYDYFPDAAKMIPKCSTQLKAVTAHFQTHTTPI<br>LLSNNFIEPLEITKEIYDLNNPEKEPKKFQTAYAKKTGDQKGYREALCKWIDFTRDFLSKYT<br>KTTSIDLSSLRPSSQYKDLGEYYAELNPLLYHISFQRIAEKEIMDAVETGKLYLFQIYNKDFA<br>KGHHGKPNLHTLYWTGLFSPENLAKTSIKLNGQAELFYRPKSRMKRMAHRLGEKMLNKK<br>LKDQKTPIPDTLYQELYDYVNHRLSHDLSDEARALLPNVITKEVSHEIIKDRRFTSDKFFFHV<br>PITLNYQAANSPSKFNQRVNAYLKEHPETPIIGIDRGERNLIYITVIDSTGKILEQRSLNTIQQF<br>DYQKKLDNREKERVAARQAWSVVGTIKDLKQGYLSQVIHEIVDLMIHYQAVVVLENLNFG<br>FKSKRTGIAEKAVYQQFEKMLIDKLNCLVLKDYPAEKVGGVLNPYQLTDQFTSFAKMGTQ<br>SGFLFYVPAPYTSKIDPLTGFVDPFVWKTIKNHESRKHFLEGFDFLHYDVKTGDFILHFKMN<br>RNLSFQRGLPGFMPAWDIVFEKNETQFDAKGTPFIAGKRIVPVIENHRFTGRYRDLYPANEL<br>IALLEEKGIVFRDGSNILPKLLENDDSHAIDTMVALIRSVLQMRNSNAATGEDYINSPVRDL<br>NGVCFDSRFQNPEWPMDADANGAYHIALKGQLLLNHLKESKDLKLQNGISNQDWLAYIQE<br>LRN |
| Cas12i1 | MKIEEGKGHHHHHHMSNKEKNASETRKAYTTKMIPRSHDRMKLLGNFMDYLMDGTPIFF<br>ELWNQFGGGIDRDIISGTANKDKISDDLLLAVNWFKVMPINSKPQGVSPSNLANLFQQYSG<br>SEPDIQAQEYFASNFDTEKHQWKDMRVEYERLLAELQLSRSDMHHDLKLMYKEKCIGLSL<br>STAHYITSVMFGTGAKNNRQTKHQFYSKVIQLLEESTQINSVEQLASIILKAGDCDSYRKLRI<br>RCSRKGATPSILKIVQDYELGTNHDDEVNVPSLIANLKEKLGRFEYECEWKCMEKIKAFLAS<br>KVGPYYLGSYSAMLENALSPIKGMTTKNCKFVLKQIDAKNDIKYENEPFGKIVEGFFDSPYF<br>ESDTNVKWVLHPHHIGESNIKTLWEDLNAIHSKYEEDIASLSEDKKEKRIKVYQGDVCQTIN<br>TYCEEVGKEAKTPLVQLLRYLYSRKDDIAVDKIIDGITFLSKKHKVEKQKINPVIQKYPSFNF<br>GNNSKLLGKIISPKDKLKHNLKCNRNQVDNYIWIEIKVLNTKTMRWEKHHYALSSTRFLEE<br>VYYPATSENPPDALAARFRTKTNGYEGKPALSAEQIEQIRSAPVGLRKVKKRQMRLEAARQ<br>QNLLPRYTWGKDFNINICKRGNNFEVTLATKVKKKKEKNYKVVLGYDANIVRKNTYAAIE<br>AHANGDGVIDYNDLPVKPIESGFVTVESQVRDKSYDQLSYNGVKLLYCKPHVESRRSFLEK<br>YRNGTMKDNRGNNIQIDFMKDFEAIADDETSLYYFNMKYCKLLQSSIRNHSSQAKEYREEI<br>FELLRDGKLSVLKLSSLSNLSFVMFKVAKSLIGTYFGHLLKKPKNSKSDVKAPPITDEDKQK<br>ADPEMFALRLALEEKRLNKVKSKKEVIANKIVAKALELRDKYGPVLIKGENISDTTKKGKK<br>SSTNSFLMDWLARGVANKVKEMVMMHQGLEFVEVNPNFTSHQDPFVHKNPENTFRARYS<br>RCTPSELTEKNRKEILSFLSDKPSKRPTNAYYNEGAMAFLATYGLKKNDVLGVSLEKFKQI<br>MANILHQRSEDQLLFPSRGGMFYLATYKLDADATSVNWNGKQFWVCNADLVAAYNVGL<br>VDIQKDFKKK |

**Table S2.** DNA and RNA sequences used to detect miRNA with Cas12i1 and AsCas12a.

| Name | Sequence |
|---|---|
| GFP Target Sense Strand | AGCACCCAGTCCGCCCTGAG |
| GFP Target Antisense Strand | CTCAGGGCGGACTGGGTGCT |
| Ca12i1 crGFP | rArArUrUrUrUrGrUrGrCrCrCrArUrCrGrUrUrGrGrCrArCrCrUrCrArGrGrGrCrGrGrArCrUrGrGrGrUrGrCrU |
| Cas12i crGFP -5 scaf | rUrUrGrUrGrCrCrCrArUrCrGrUrUrGrGrCrArCrCrUrCrArGrGrGrCrGrGrArCrUrGrGrGrUrGrCrU |
| Cas12i crGFP -10 scaf | rCrCrCrArUrCrGrUrUrGrGrCrArCrCrUrCrArGrGrGrCrGrGrArCrUrGrGrGrUrGrCrU |
| Cas12i crGFP -14 scaf | rUrCrGrUrUrGrGrCrArCrCrUrCrArGrGrGrCrGrGrArCrUrGrGrGrUrGrCrU |
| Cas12i GFP Spacer Only | rCrUrCrArGrGrGrCrGrGrArCrUrGrGrGrUrGrCrU |
| miR-21 20nt cDNA | ACAGCCCATCGACTGGTGTT |
| miR-122 20nt cDNA | TATTTAGTGTGATAATGGCG |
| miR-155 20nt cDNA | ACCCCTATCACGATTAGCAT |
| miR-21 RNA Target | rCrArArCrArCrCrArGrUrCrGrArUrGrGrGrCrUrGrU |
| miR-122 RNA Target | rArArCrGrCrCrArUrUrArUrCrArCrArCrUrArArArUrA |
| miR-155 RNA Target | rUrUrArArUrGrCrUrArArUrCrGrUrGrArUrArGrGrGrGrU |
| miR-21 20 nt 3' Handle Cas12i1 (ΨDNA) | ACAGCCCATCGACTGGTGTTAATTTTTGTGCCCATCGTTGGCAC |
| miR-21 20 nt 5' Handle Cas12i1 | AATTTTTGTGCCCATCGTTGGCACACAGCCCATCGACTGGTGTT |
| miR-21 20 nt double handle Cas12i1 | AATTTTTGTGCCCATCGTTGGCACACAGCCCATCGACTGGTGTTAATTTTTGTGCCCATCGTTGGCAC |
| miR-21 ΨDNA AsCas12a | ACAGCCCATCGACTGGTGTTTAGATGTGAATCATCTTTAAT |
| miR-122 ΨDNA AsCas12a | TATTTAGTGTGATAATGGCGTAGATGTGAATCATCTTTAAT |
| miR-155 ΨDNA AsCas12a | ACCCCTATCACGATTAGCATTAGATGTGAATCATCTTTAAT |
| miR-122 ΨDNA Cas12i1 | TATTTAGTGTGATAATGGCGAATTTTTGTGCCCATCGTTGGCAC |
| miR-155 ΨDNA Cas12i1 | ACCCCTATCACGATTAGCATAATTTTTGTGCCCATCGTTGGCAC |
| FQ Reporter (ssDNA) | FAM/TTATT/Iowa Black |
| FQ ssRNA | FAM/rUrUrUrUrU/Iowa Black |

**Table S3.** DNA and RNA sequences used for EMSA assay.

| Name | Sequence |
|---|---|
| ΨDNA AsCas12a | AGCACCCAGTCCGCCCTGAGTAGATGTGAATCATCTTTAAT |
| ΨDNA Cas12i1 | AGCACCCAGTCCGCCCTGAGAATTTTTGTGCCCATCGTTGGCAC |
| RNA Target | rCrUrCrArGrGrGrCrGrGrArCrUrGrGrGrUrGrCrU |
| crRNA AsCas12a | rUrArArUrUrUrCrUrArCrUrArArGrUrGrUrArGrArUrCrUrCrArGrGrGrCrGrGrArCr UrGrGrGrUrGrCrU |
| crRNA Cas12i1 | rArArUrUrUrUrUrGrUrGrCrCrCrArUrCrGrUrUrGrGrCrArCrCrUrCrArGrGrGrCrGr GrArCrUrGrGrGrUrGrCrU |
| ssDNA Target | AGCACCCAGTCCGCCCTGAG |

**Table S4.** DNA and RNA sequences used for BLI assay.

| Name | Sequence |
|---|---|
| AsCas12a crGFP w/ biotin | rUrArArUrUrUrCrUrArCrUrArArGrUrGrUrArGrArUrCrUrCrArGrGrGrCrGrGrArCrU rGrGrGrUrGrCrU/3Bio/ |
| Cas12i1 crGFP w/ biotin | rArArUrUrUrUrUrGrUrGrCrCrCrArUrCrGrUrUrGrGrCrArCrCrUrCrArGrGrGrCrGrG rArCrUrGrGrGrUrGrCrU/3Bio/ |
| GFP RNA spacer w/ biotin | rCrUrCrArGrGrGrCrGrGrArCrUrGrGrGrUrGrCrU/3Bio/ |
| GFP cDNA w/ biotin | AGCACCCAGTCCGCCCTGAG/3Bio/ |
| AsCas12a GFP cDNA 5' handle w/ biotin | TAGATGTGAATCATCTTTAATAGCACCCAGTCCGCCCTGAG/3Bio/ |
| AsCas12a GFP ΨDNA w/ biotin | AGCACCCAGTCCGCCCTGAGTAGATGTGAATCATCTTTAAT/3Bio/ |
| AsCas12a GFP cDNA double handle w/ biotin | TAGATGTGAATCATCTTTAATAGCACCCAGTCCGCCCTGAGTAGATGTGAAT CATCTTTAAT/3Bio/ |
| Cas12i1 GFP cDNA 5' handle w/ biotin | AATTTTTGTGCCCATCGTTGGCACAGCACCCAGTCCGCCCTGAG/3Bio/ |
| Cas12i1 GFP ΨDNA w/ biotin | AGCACCCAGTCCGCCCTGAGAATTTTTGTGCCCATCGTTGGCAC/3Bio/ |
| Cas12i1 GFP cDNA double handle w/ biotin | AATTTTTGTGCCCATCGTTGGCACAGCACCCAGTCCGCCCTGAGAATTTTTGT GCCCATCGTTGGCAC/3Bio/ |

**Table S5.** DNA and RNA sequences for RNA cis-cleavage.

| Name | Sequence |
| --- | --- |
| Target | FAM/rArGrArCrCrGrUrGrCrArUrCrArUrCrCrUrArArArCrCrUrCrGrUrCrCrGrCrCrC rUrGrArGrGrArArCrGrCrUrGrArArGrCrGrCrCrUrUrGrUrGrGrUrArC/Cy5 |
| AsCas12a ΨDNA | TCAGCGTTCCTCGGGCGGAC TAGATGTGAATCATCTTTAAT |
| Cas12i1 ΨDNA | TCAGCGTTCCTCGGGCGGAC AATTTTTGTGCCCATCGTTGGCAC |
| DNAzyme 10-23 | GAGGTTTAGGA GGCTAGCTACAACGA GATGCACGG |

**Table S6.** DNA and RNA sequences used for long RNA detection with synthethic HIV RNA fragments

| Name | Sequences |
|---|---|
| T7-HIV Geneblock 1 - DNA fragment with T7 promoter used to produce a fragment of genomic HIV RNA through in vitro transcription with T7 RNA Pol | TAATACGACTCACTATAGAAAAGATGGATAATCCTGGGATTAAATAAAATAG<br>TAAGAATGTATAGCCCTACCAGCATTCTGGACATAAGACAAGGACCAAAGGA<br>ACCCTTTAGAGACTATGTAGACCGGTTCTATAAAACTCTAAGAGCCGAGCAA<br>GCTTCACAGGAGGTAAAAAATTGGATGACAGAAACCTTGTTGGTCCAAAATG<br>CGAACCCAGATTGTAAGACTATTTTAAAAGCATTGGGACCAGCGGCTACACT<br>AGAAGAAATGATGACAGCATGTCAGGGAGTAGGAGGACCCGGCCATAAGGC<br>AAGAGTTTTGGCTGAAGCAATGAGCCAAGTAACAAATTCAGCTACCATAATG<br>ATGCAGAGAGGCAATTTTAGGAACCAAAGAAAGATTGTTAAGTGTTTCAATT<br>GTGGCAAAGAAGGGCACACAGCCAGAAATTGCAGGGCCCCTAGGAAAAAGG<br>GCTGTTGGAAATGTGGAAAGGAAGGACACCAAATGAAAGATTGTACTGAGAG<br>ACAGGCTAATTTTTTAGGGAAGATCTGGCCTTCCTACAAGGGAAGGCCAGGG<br>AATTTTCTTCAGAGCAGACCAGAGCCAACAGCCCCACCAGAAGAGAGCTTCA<br>GGTCTGGGGTAGAGACAACAACTCCCCCTCAGAAGCAGGAGCCGATAGACAA<br>GGAACTGTATCCTTTAACTTCCCTCAGGTCACTCTTTGGCAACGACCCCTCGTC<br>ACAATAAAGATAGGGGGGCAACTAAAGGAAGCTCTATTAGATACAGGAGCA<br>GATGATACAGTATTAGAAGAAATGAGTTTGCCAGGAAGATGGAAACCAAAAA<br>TGATAGGGGGAATTGGAGGTTTTATCAAAGTAAGACAGTATGATCAGATACT<br>CATAGAAATCTGTGGACATAAAGCTATAGGTACAGTATTAGTAGGACCTACA<br>CCTGTCAACATAATTGGAAGAAATCTGTTGACTCAGATTGGTTGCACTTTAAA<br>TTTTCCCATTAGCCCTATTGAGACTGTACCAGTAAAATTAAAGCCAGGAATGG<br>ATGGCCCAAAAGTTAAACAATGGCCATTGACAGAAGAAAAAATAAAAGCATT<br>AGTAGAAATTTGTACAGAGATGGAAAAGGAAGGGAAAATTTCAAAAATTGGG<br>CCTGAAAATCCATACAATACTCCAGTATTTGCCATAAAGAAAAAAGACAGTA<br>CTAAATGGAGAAAATTAGTAGATTTCAGAGAACTTAATAAGAGAACTCAAGA<br>CTTCTGGGAAGTTCAATTAGGAATACCACATCCCGCAGGGTTAAAAAAGAAA<br>AAATCAGTAACAGTACTGGATGTGGGTGATGCATATTTTTCAGTTCCCTTAGA<br>TGAAGACTTCAGGAAGTATACTGCATTTACCATACCTAGTATAAACAATGAGA<br>CACCAGGGATTAGATATCAGTACAATGTGCTTCCACAGGGATGGAAAGGATC<br>ACCAGCAATATTCCAAAGTAGCATGACAAAAATCTTAGAGCCTTTTAGAAAA<br>CAAAATCCAGACATAGTTATCTATCAATACATGGATGATTTGTATGTAGGATC<br>TGACTTAGAAATAGGGCAGCATAGAACAAAAATAGAGGAGCTGAGACAACA<br>TCTGTTGAGGTGGGGACTTACCACACCAGACAAAAAACATCAGAAAGAACCT<br>CCATTCCTTTGGATGGGTTATGAACTCCATCCTGATAAATGGACAGTACAGCC<br>TATAGTGCTGCCAGAAAAAGACAGCTGGACTGTCAATGACATACAGAA |

| | |
|---|---|
| T7-HIV Geneblock 2 - DNA fragment with T7 promoter used to produce a fragment of genomic HIV RNA through in vitro transcription with T7 RNA Pol | TAATACGACTCACTATAGGGGCGGGAATCAAGCAGGAATTTGGAATTCCCTA<br>CAATCCCCAAAGTCAAGGAGTAGTAGAATCTATGAATAAAGAATTAAAGAAA<br>ATTATAGGACAGGTAAGAGATCAGGCTGAACATCTTAAGACAGCAGTACAAA<br>TGGCAGTATTCATCCACAATTTTAAAAGAAAAGGGGGGATTGGGGGGTACAG<br>TGCAGGGGAAAGAATAGTAGACATAATAGCAACAGACATACAAACTAAAGA<br>ATTACAAAAACAAATTACAAAAATTCAAAATTTTCGGGTTTATTACAGGGACA<br>GCAGAAATCCACTTTGGAAAGGACCAGCAAAGCTCCTCTGGAAAGGTGAAGG<br>GGCAGTAGTAATACAAGATAATAGTGACATAAAAGTAGTGCCAAGAAGAAA<br>AGCAAAGATCATTAGGGATTATGGAAAACAGATGGCAGGTGATGATTGTGTG<br>GCAAGTAGACAGGATGAGGATTAGAACATGGAAAAGTTTAGTAAAACACCAT<br>ATGTATGTTTCAGGGAAAGCTAGGGGATGGTTTTATAGACATCACTATGAAAG<br>CCCTCATCCAAGAATAAGTTCAGAAGTACACATCCCACTAGGGGATGCTAGA<br>TTGGTAATAACAACATATTGGGGTCTGCATACAGGAGAAAGAGACTGGCATT<br>TGGGTCAGGGAGTCTCCATAGAATGGAGGAAAAAGAGATATAGCACACAAGT<br>AGACCCTGAACTAGCAGACCAACTAATTCATCTGTATTACTTTGACTGTTTTTC<br>AGACTCTGCTATAAGAAAGGCCTTATTAGGACACATAGTTAGCCCTAGGTGTG<br>AATATCAAGCAGGACATAACAAGGTAGGATCTCTACAATACTTGGCACTAGC<br>AGCATTAATAACACCAAAAAAGATAAAGCCACCTTTGCCTAGTGTTACGAAA<br>CTGACAGAGGATAGATGGAACAAGCCCCAGAAGACCAAGGGCCACAGAGGG<br>AGCCACACAATGAATGGACACTAGAGCTTTTAGAGGAGCTTAAGAATGAAGC<br>TGTTAGACATTTTCCTAGGATTTGGCTCCATGGCTTAGGGCAACATATCTATG<br>AAACTTATGGGGATACTTGGGCAGGAGTGGAAGCCATAATAAGAATTCTGCA<br>ACAACTGCTGTTTATCCATTTTCAGAATTGGGTGTCGACATAGCAGAATAGGC<br>GTTACTCGACAGAGGAGAGCAAGAAATGGAGCCAGTAGATCCTAGACTAGAG<br>CCCTGGAAGCATCCAGGAAGTCAGCCTAAAACTGCTTGTACCAATTGCTATTG<br>TAAAAAGTGTTGCTTTCATTGCCAAGTTTGTTTCATAACAAAAGCCTTAGGCA<br>TCTCCTATGGCAGGAAGAAGCGGAGACAGCGACGAAGAGCTCATCAGAACAG<br>TCAGACTCATCAAGCTTCTCTATCAAAGCAGTAAGTAGTACATGTAATGCAAC<br>CTATACCAATAGTAGCAATAGTAGCATTAGTAGTAGCAATAATAATAGCAAT<br>AGTTGTGTGGTCCATAGTAATCATAGAATATAGGAAAATATTAAGACAAAGA<br>AAAATAGACAGGTTAATTGATAGACTAATAGAAAGAGCAGAAGACAGTGGC<br>AATGAGAGTGAAGGAGAAATATCAGCACTTGTGGAGATGGGGGTGGAGATG<br>GGGCACCATGCTCCTTGGGATGTTGATGATCTGTAGTGCTACAGAAAAATTGT<br>GG |

| | |
|---|---|
| T7-HIV Geneblock 3 - DNA fragment with T7 promoter used to produce a fragment of genomic HIV RNA through in vitro transcription with T7 RNA Pol | TAATACGACTCACTATAGAGACCCAACAACAATACAAGAAAAAGAATCCGTA<br>TCCAGAGAGGACCAGGGAGAGCATTTGTTACAATAGGAAAAATAGGAAATAT<br>GAGACAAGCACATTGTAACATTAGTAGAGCAAAATGGAATAACACTTTAAAA<br>CAGATAGCTAGCAAATTAAGAGAACAATTTGGAAATAATAAAACAATAATCT<br>TTAAGCAATCCTCAGGAGGGGACCCAGAAATTGTAACGCACAGTTTTAATTGT<br>GGAGGGGAATTTTTCTACTGTAATTCAACACAACTGTTTAATAGTACTTGGTT<br>TAATAGTACTTGGAGTACTGAAGGGTCAAATAACACTGAAGGAAGTGACACA<br>ATCACCCTCCCATGCAGAATAAAACAAATTATAAACATGTGGCAGAAAGTAG<br>GAAAAGCAATGTATGCCCCTCCCATCAGTGGACAAATTAGATGTTCATCAAAT<br>ATTACAGGGCTGCTATTAACAAGAGATGGTGGTAATAGCAACAATGAGTCCG<br>AGATCTTCAGACCTGGAGGAGGAGATATGAGGGACAATTGGAGAAGTGAATT<br>ATATAAATATAAAGTAGTAAAAATTGAACCATTAGGAGTAGCACCCACCAAG<br>GCAAAGAGAAGAGTGGTGCAGAGAGAAAAAAGAGCAGTGGGAATAGGAGCT<br>TTGTTCCTTGGGTTCTTGGGAGCAGCAGGAAGCACTATGGGCGCAGCCTCAAT<br>GACGCTGACGGTACAGGCCAGACAATTATTGTCTGGTATAGTGCAGCAGCAG<br>AACAATTTGCTGAGGGCTATTGAGGCGCAACAGCATCTGTTGCAACTCACAGT<br>CTGGGGCATCAAGCAGCTCCAGGCAAGAATCCTGGCTGTGGAAAGATACCTA<br>AAGGATCAACAGCTCCTGGGGATTTGGGGTTGCTCTGGAAAACTCATTTGCAC<br>CACTGCTGTGCCTTGGAATGCTAGTTGGAGTAATAAATCTCTGGAACAGATTT<br>GGAATCACACGACCTGGATGGAGTGGGACAGAGAAATTAACAATTACACAAG<br>CTTAATACACTCCTTAATTGAAGAATCGCAAAACCAGCAAGAAAAGAATGAA<br>CAAGAATTATTGGAATTAGATAAATGGGCAAGTTTGTGGAATTGGTTTAACAT<br>AACAAATTGGCTGTGGTATATAAAATTATTCATAATGATAGTAGGAGGCTTGG<br>TAGGTTTAAGAATAGTTTTTGCTGTACTTTCTATAGTGAATAGAGTTAGGCAG<br>GGATATTCACCATTATCGTTTCAGACCCACCTCCCAACCCCGAGGGGACCCGA<br>CAGGCCCGAAGGAATAGAAGAAGAAGGTGGAGAGAGAGACAGAGACAGATC<br>CATTCGATTAGTGAACGGATCCTTGGCACTTATCTGGGACGATCTGCGGAGCC<br>TGTGCCTCTTCAGCTACCACCGCTTGAGAGACTTACTCTTGATTGTAACGAGG<br>ATTGTGGAACTTCTGGGACGCAGGGGGTGGGAAGCCCTCAAATATTGGTGGA<br>ATCTCCTACAGTATTGGAGTCAGGAACTAAAGAATAGTGCTGTTAGCTTGCTC<br>AATGCCACAGCCATAGCAGTAGCTGAGGGGACAGATAGGGTTATAGAAGTAG<br>TACAAGGAGCTTGTAGAGCTATTCGCCACATACCTAGAAGAATAAGACAGGG<br>C |
| HIV ΨDNA Cas12i1_1 | CTG GTC TAA CCA GAG AGA CCA ATT TTT GTG CCC ATC GTT GGC AC |
| HIV ΨDNA Cas12i1_2 | TTG AAG CAC TCA AGG CAA GCA ATT TTT GTG CCC ATC GTT GGC AC |
| HIV ΨDNA Cas12i1_3 | TGG CGT ACT CAC CAG TCG CCA ATT TTT GTG CCC ATC GTT GGC AC |
| HIV ΨDNA Cas12i1_4 | TCC CAG TAT TTG TCT ACA GCA ATT TTT GTG CCC ATC GTT GGC AC |
| HIV ΨDNA Cas12i1_5 | CTG ACC TGA TTG CTG TGT CCA ATT TTT GTG CCC ATC GTT GGC AC |
| HIV ΨDNA Cas12i1_6 | GCT TCC TCA TTG ATG GTC TCA ATT TTT GTG CCC ATC GTT GGC AC |
| HIV ΨDNA Cas12i1_7 | GTC CAG AAT GCT GGT AGG GCA ATT TTT GTG CCC ATC GTT GGC AC |
| HIV ΨDNA Cas12i1_8 | GGT CCT CCT ACT CCC TGA CAA ATT TTT GTG CCC ATC GTT GGC AC |
| HIV ΨDNA Cas12i1_9 | ATT AGC CTG TCT CTC AGT ACA ATT TTT GTG CCC ATC GTT GGC AC |
| HIV ΨDNA Cas12i1_10 | TTG CCA AAG AGT GAC CTG AGA ATT TTT GTG CCC ATC GTT GGC AC |
| HIV ΨDNA Cas12i1_11 | TTG ACA GGT GTA GGT CCT ACA ATT TTT GTG CCC ATC GTT GGC AC |

| | |
|---|---|
| HIV ΨDNA Cas12i1_12 | TCT TCT GTC AAT GGC CAT TGA ATT TTT GTG CCC ATC GTT GGC AC |
| HIV ΨDNA Cas12i1_13 | CCC AGA AGT CTT GAG TTC TCA ATT TTT GTG CCC ATC GTT GGC AC |
| HIV ΨDNA Cas12i1_14 | TGG AAT ATT GCT GGT GAT CCA ATT TTT GTG CCC ATC GTT GGC AC |
| HIV ΨDNA Cas12i1_15 | TGG CAG CAC TAT AGG CTG TAA ATT TTT GTG CCC ATC GTT GGC AC |
| HIV ΨDNA Cas12i1_16 | TGC CAG TTC TAG CTC TGC TTA ATT TTT GTG CCC ATC GTT GGC AC |
| HIV ΨDNA Cas12i1_17 | GTT TCC TTT TGT ATG GGC AGA ATT TTT GTG CCC ATC GTT GGC AC |
| HIV ΨDNA Cas12i1_18 | TTA GTC TCC CTG TTA GCT GCA ATT TTT GTG CCC ATC GTT GGC AC |
| HIV ΨDNA Cas12i1_19 | GTA CCC ATG CCA GAT AGA CCA ATT TTT GTG CCC ATC GTT GGC AC |
| HIV ΨDNA Cas12i1_20 | ACA GTC TAC TTG TCC ATG CAA ATT TTT GTG CCC ATC GTT GGC AC |
| HIV ΨDNA Cas12i1_21 | TGC TGC CAT TGT CAG TAT GTA ATT TTT GTG CCC ATC GTT GGC AC |
| HIV ΨDNA Cas12i1_22 | TTC TTT CCC CTG CAC TGT ACA ATT TTT GTG CCC ATC GTT GGC AC |
| HIV ΨDNA Cas12i1_23 | CAA TCA TCA CCT GCC ATC TGA ATT TTT GTG CCC ATC GTT GGC AC |
| HIV ΨDNA Cas12i1_24 | TGG GAT GTG TAC TTC TGA ACA ATT TTT GTG CCC ATC GTT GGC AC |
| HIV ΨDNA Cas12i1_25 | AGT ATT GTA GAG ATC CTA CCA ATT TTT GTG CCC ATC GTT GGC AC |
| HIV ΨDNA Cas12i1_26 | CAG CTT CAT TCT TAA GCT CCA ATT TTT GTG CCC ATC GTT GGC AC |
| HIV ΨDNA Cas12i1_27 | TTC CTG GAT GCT TCC AGG GCA ATT TTT GTG CCC ATC GTT GGC AC |
| HIV ΨDNA Cas12i1_28 | ATG GAC CAC ACA ACT ATT GCA ATT TTT GTG CCC ATC GTT GGC AC |
| HIV ΨDNA Cas12i1_29 | CTG TAG CAC TAC AGA TCA TCA ATT TTT GTG CCC ATC GTT GGC AC |
| HIV ΨDNA Cas12i1_30 | GCT TTA GGC TTT GAT CCC ATA ATT TTT GTG CCC ATC GTT GGC AC |
| HIV ΨDNA Cas12i1_31 | TGA CTG AGG TGT TAC AAC TTA ATT TTT GTG CCC ATC GTT GGC AC |
| HIV ΨDNA Cas12i1_32 | TGA GTT GAT ACT ACT GGC CTA ATT TTT GTG CCC ATC GTT GGC AC |
| HIV ΨDNA Cas12i1_33 | TGT TAC AAT GTG CTT GTC TCA ATT TTT GTG CCC ATC GTT GGC AC |
| HIV ΨDNA Cas12i1_34 | GTG ATT GTG TCA CTT CCT TCA ATT TTT GTG CCC ATC GTT GGC AC |
| HIV ΨDNA Cas12i1_35 | TAT CTC CTC CTC CAG GTC TGA ATT TTT GTG CCC ATC GTT GGC AC |
| HIV ΨDNA Cas12i1_36 | TCA GCA AAT TGT TCT GCT GCA ATT TTT GTG CCC ATC GTT GGC AC |
| HIV ΨDNA Cas12i1_37 | CAT CCA GGT CGT GTG ATT CCA ATT TTT GTG CCC ATC GTT GGC AC |
| HIV ΨDNA Cas12i1_38 | ATC CCT GCC TAA CTC TAT TCA ATT TTT GTG CCC ATC GTT GGC AC |

| | |
|---|---|
| HIV ΨDNA Cas12i1_39 | AGT AAG TCT CTC AAG CGG TGA ATT TTT GTG CCC ATC GTT GGC AC |
| HIV ΨDNA Cas12i1_40 | TAT TCT TCT AGG TAT GTG GCA ATT TTT GTG CCC ATC GTT GGC AC |
| HIV ΨDNA Cas12i1_41 | CTC TTG TGC TTC TAG CCA GGA ATT TTT GTG CCC ATC GTT GGC AC |
| HIV ΨDNA Cas12i1_42 | CAA CTG GTA CTA GCT TGT AGA ATT TTT GTG CCC ATC GTT GGC AC |
| HIV ΨDNA Cas12i1_43 | AAG TCC CTT GTA GCA AGC TCA ATT TTT GTG CCC ATC GTT GGC AC |
| HIV ΨDNA Cas12i1_44 | TAT ATG CAG GAT CTG AGG GCA ATT TTT GTG CCC ATC GTT GGC AC |
| HIV ΨDNA Cas12i1_45 | GAA GCA CTC AAG GCA AGC TTA ATT TTT GTG CCC ATC GTT GGC AC |
| HIV ΨDNA Cas12i1_46 | ATC CCG AAT CCT GCA AAG CTA ATT TTT GTG CCC ATC GTT GGC AC |
| HIV ΨDNA Cas12i1_47 | TAT ATC CAC TGG CTA CAT GAA ATT TTT GTG CCC ATC GTT GGC AC |
| HIV ΨDNA Cas12i1_48 | GTA CTG TCC ATT TAT CAG GAA ATT TTT GTG CCC ATC GTT GGC AC |
| HIV ΨDNA AsCas12a_1 | CTG GTC TAA CCA GAG AGA CCT AGA TGT GAA TCA TCT TTA AT |
| HIV ΨDNA AsCas12a_2 | TTG AAG CAC TCA AGG CAA GCT AGA TGT GAA TCA TCT TTA AT |
| HIV ΨDNA AsCas12a_3 | TGG CGT ACT CAC CAG TCG CCT AGA TGT GAA TCA TCT TTA AT |
| HIV ΨDNA AsCas12a_4 | TCC CAG TAT TTG TCT ACA GCT AGA TGT GAA TCA TCT TTA AT |
| HIV ΨDNA AsCas12a_5 | CTG ACC TGA TTG CTG TGT CCT AGA TGT GAA TCA TCT TTA AT |
| HIV ΨDNA AsCas12a_6 | GCT TCC TCA TTG ATG GTC TCT AGA TGT GAA TCA TCT TTA AT |
| HIV ΨDNA AsCas12a_7 | GTC CAG AAT GCT GGT AGG GCT AGA TGT GAA TCA TCT TTA AT |
| HIV ΨDNA AsCas12a_8 | GGT CCT CCT ACT CCC TGA CAT AGA TGT GAA TCA TCT TTA AT |
| HIV ΨDNA AsCas12a_9 | ATT AGC CTG TCT CTC AGT ACT AGA TGT GAA TCA TCT TTA AT |
| HIV ΨDNA AsCas12a_10 | TTG CCA AAG AGT GAC CTG AGT AGA TGT GAA TCA TCT TTA AT |
| HIV ΨDNA AsCas12a_11 | TTG ACA GGT GTA GGT CCT ACT AGA TGT GAA TCA TCT TTA AT |
| HIV ΨDNA AsCas12a_12 | TCT TCT GTC AAT GGC CAT TGT AGA TGT GAA TCA TCT TTA AT |
| HIV ΨDNA AsCas12a_13 | CCC AGA AGT CTT GAG TTC TCT AGA TGT GAA TCA TCT TTA AT |
| HIV ΨDNA AsCas12a_14 | TGG AAT ATT GCT GGT GAT CCT AGA TGT GAA TCA TCT TTA AT |
| HIV ΨDNA AsCas12a_15 | TGG CAG CAC TAT AGG CTG TAT AGA TGT GAA TCA TCT TTA AT |
| HIV ΨDNA AsCas12a_16 | TGC CAG TTC TAG CTC TGC TTT AGA TGT GAA TCA TCT TTA AT |
| HIV ΨDNA AsCas12a_17 | GTT TCC TTT TGT ATG GGC AGT AGA TGT GAA TCA TCT TTA AT |
| HIV ΨDNA AsCas12a_18 | TTA GTC TCC CTG TTA GCT GCT AGA TGT GAA TCA TCT TTA AT |
| HIV ΨDNA AsCas12a_19 | GTA CCC ATG CCA GAT AGA CCT AGA TGT GAA TCA TCT TTA AT |
| HIV ΨDNA AsCas12a_20 | ACA GTC TAC TTG TCC ATG CAT AGA TGT GAA TCA TCT TTA AT |
| HIV ΨDNA AsCas12a_21 | TGC TGC CAT TGT CAG TAT GTT AGA TGT GAA TCA TCT TTA AT |
| HIV ΨDNA AsCas12a_22 | TTC TTT CCC CTG CAC TGT ACT AGA TGT GAA TCA TCT TTA AT |
| HIV ΨDNA AsCas12a_23 | CAA TCA TCA CCT GCC ATC TGT AGA TGT GAA TCA TCT TTA AT |
| HIV ΨDNA AsCas12a_24 | TGG GAT GTG TAC TTC TGA ACT AGA TGT GAA TCA TCT TTA AT |
| HIV ΨDNA AsCas12a_25 | AGT ATT GTA GAG ATC CTA CCT AGA TGT GAA TCA TCT TTA AT |
| HIV ΨDNA AsCas12a_26 | CAG CTT CAT TCT TAA GCT CCT AGA TGT GAA TCA TCT TTA AT |

| | |
|---|---|
| HIV ΨDNA AsCas12a_27 | TTC CTG GAT GCT TCC AGG GCT AGA TGT GAA TCA TCT TTA AT |
| HIV ΨDNA AsCas12a_28 | ATG GAC CAC ACA ACT ATT GCT AGA TGT GAA TCA TCT TTA AT |
| HIV ΨDNA AsCas12a_29 | CTG TAG CAC TAC AGA TCA TCT AGA TGT GAA TCA TCT TTA AT |
| HIV ΨDNA AsCas12a_30 | GCT TTA GGC TTT GAT CCC ATT AGA TGT GAA TCA TCT TTA AT |
| HIV ΨDNA AsCas12a_31 | TGA CTG AGG TGT TAC AAC TTT AGA TGT GAA TCA TCT TTA AT |
| HIV ΨDNA AsCas12a_32 | TGA GTT GAT ACT ACT GGC CTT AGA TGT GAA TCA TCT TTA AT |
| HIV ΨDNA AsCas12a_33 | TGT TAC AAT GTG CTT GTC TCT AGA TGT GAA TCA TCT TTA AT |
| HIV ΨDNA AsCas12a_34 | GTG ATT GTG TCA CTT CCT TCT AGA TGT GAA TCA TCT TTA AT |
| HIV ΨDNA AsCas12a_35 | TAT CTC CTC CTC CAG GTC TGT AGA TGT GAA TCA TCT TTA AT |
| HIV ΨDNA AsCas12a_36 | TCA GCA AAT TGT TCT GCT GCT AGA TGT GAA TCA TCT TTA AT |
| HIV ΨDNA AsCas12a_37 | CAT CCA GGT CGT GTG ATT CCT AGA TGT GAA TCA TCT TTA AT |
| HIV ΨDNA AsCas12a_38 | ATC CCT GCC TAA CTC TAT TCT AGA TGT GAA TCA TCT TTA AT |
| HIV ΨDNA AsCas12a_39 | AGT AAG TCT CTC AAG CGG TGT AGA TGT GAA TCA TCT TTA AT |
| HIV ΨDNA AsCas12a_40 | TAT TCT TCT AGG TAT GTG GCT AGA TGT GAA TCA TCT TTA AT |
| HIV ΨDNA AsCas12a_41 | CTC TTG TGC TTC TAG CCA GGT AGA TGT GAA TCA TCT TTA AT |
| HIV ΨDNA AsCas12a_42 | CAA CTG GTA CTA GCT TGT AGT AGA TGT GAA TCA TCT TTA AT |
| HIV ΨDNA AsCas12a_43 | AAG TCC CTT GTA GCA AGC TCT AGA TGT GAA TCA TCT TTA AT |
| HIV ΨDNA AsCas12a_44 | TAT ATG CAG GAT CTG AGG GCT AGA TGT GAA TCA TCT TTA AT |
| HIV ΨDNA AsCas12a_45 | GAA GCA CTC AAG GCA AGC TTT AGA TGT GAA TCA TCT TTA AT |
| HIV ΨDNA AsCas12a_46 | ATC CCG AAT CCT GCA AAG CTT AGA TGT GAA TCA TCT TTA AT |
| HIV ΨDNA AsCas12a_47 | TAT ATC CAC TGG CTA CAT GAT AGA TGT GAA TCA TCT TTA AT |
| HIV ΨDNA AsCas12a_48 | GTA CTG TCC ATT TAT CAG GAT AGA TGT GAA TCA TCT TTA AT |

.

**Table S7.** DNA scaffold sequences from diverse CRISPR-Cas systems for ΨDNA-medited RNA detection

| CRISPR system | Organism | Predicted Mature Repeat |
|---|---|---|
| Cas12a | Francisella hispaniensis FSC454 | AAATTATTTAAAGTTCTTAGAC |
| | Eubacterium eligens | AAATTATTTAAGGTTATTCAAAC |
| | Candidatus Moranbacteria bacterium | AAATTGTGTAGGTCTTATTGCG |
| | Moraxella bovoculi | AAATTTCTACTGTTTGTAGAT |
| | Succinivibrio sp. | AAATTTCTACTTATGTAGAT |
| | Candidatus Peregrinibacteria bacterium | AATCCTATAGGTCGTTTAGAG |
| | Prevotella bryantii B14 | AATTAAATAAGCTTTATAGCC |
| | Helcococcus kunzii | AATTAATAGTATCTCTTAAAG |
| | Muribaculaceae bacterium | AATTACAGGCTTTATGTAGCC |
| | Prevotellamassilia sp. | AATTATAAAGGCATTATAGCC |
| | Butyrivibrio hungatei | AATTATCTTTAAGTCTTAGAC |
| | Pseudobutyrivibrio xylanivorans | AATTATTTAAAGGTTCTAAGC |
| | Francisella cf. novicida Fx1 | AATTATTTAAAGTTCTTAGAC |
| | Candidatus Methanomethylophilus alvus | AATTCTGAATGAGTTTTAGAC |
| | Flavobacterium jumunjinense | AATTTAGTTTGTCTTTAAAAC |
| | Sedimentisphaera cyanobacteriorum L21-RPul-D3 | AATTTTATAAGGCCTTTAGAC |
| | Sneathia amnii (fusobacteria) SN35 | AATTTTATTTGGGTTCTAAAC |
| | Alicyclobacillus acidoterrestris | AGCGATCTGAGAAGTGGCAC |
| | Candidatus Methanoplasma termitum (euryarchaeotes) MpT1 | GAATCTCTACTCTTTGTAGAT |
| | Eubacterium rectale M104/1 (firmicutes) | GAATGTCTACTGGGGTAGATC |
| | Lachnospiraceae bacterium GAM79 | GAATTTCTACTAGTGTAGAT |
| | Ruminococcus sp. JE7A12 (firmicutes) | GAATTTCTACTATTGTAGAT |
| | Moraxella bovoculi | TAAATTTCTACTGTTTGTAGAT |
| | Lachnospiraceae bacterium ND2006 | TAATTTCTACTAAGTGTAGAT |
| | Prevotella bryantii | TAATTTCTACTATTGTAGAT |
| | Acidaminoscoccus Cas12a | TAATTTCTACTCTTGTAGAT |

| | | |
|---|---|---|
| | Francisella tularensis subsp. novicida F6168 (g-proteobacteria) | TAATTTCTACTGTTGTAGAT |
| | Lachnospira eligens (firmicutes) | TAATTTCTACTTTGTAGAT |
| | Catenovulum sp. CCB-QB4 (g-proteobacteria) | TCACCTACGTAACCGTGAACT |
| | PdCas12a | TAATTTCTACTTCGGTAGAT |
| | LpCas12a | TAATTTCTACTGTGTGTAGAT |
| | Methylobacterium nodulans ORS 2060 | AGGATCTGGCGCCCACTGCGAC |
| | Akkermansiaceae bacterium | CACATCGCGGGAACTGCGGC |
| | Pelobacter propionicus DSM 2379 | CCCCGCGCATGCGGGGAACAC |
| | Bacillus sp. NSP2.1 | CCTCGGGTCCTCAATGTAAC |
| | Alicyclobacillus acidoterrestris | CGAGCGATCTGAGAAGTGGCAC |
| | Brevibacillus agri (firmicutes) DSM 6348 | CTTTCCACTAAGCTTTCGAAC |
| | Brevibacillus sp. FJAT-54423 | GAAAAGCTGAGAAGTTAGCAC |
| | Sulfobacillus thermotolerans (firmicutes) | GAATGCTTAGGTTGTTGGCAC |
| | Phycisphaeraceae bacterium | GCCGTGTTGGCCGATGCGGC |
| | Methylacidimicrobium sp. AP8 | GGCCTGCGCCGACCCGAACCGC |
| | Phycisphaeraceae bacterium | GGGCCGCGTCGGCCTCCGCGGC |
| | Opitutaceae bacterium TAV5 | TGAAACGGCATTCGCTGCGGC |
| | Sulfobacillus thermotolerans | TGCTTAGGTTGTTGGCAC |
| | Brevibacillus sp. HD3.3A | TGGAAAGCTTCGGGATTAGCAC |
| | Brevibacillus sp. FJAT-54423 | TGGAGTGCGTGGATTGAAAC |
| Other Cas12 & Racr | Cas12d | ACCCGTAAAGCAGAGCGATGAAGGC |
| | PlmCas12e | TATTTATCGGAGATATCTTCAAAC |
| | Cas12f | CGCGCCCCTGATGAATGGACAC |
| | Cas12h1 | GCTAGAGGGAGGTCAGAGCAC |
| | Cas12i1 | AATTTTTGTGCCCATCGTTGGCAC |
| | Cas12i2 | AGAAATCCGTCTTTCATTGACGG |
| | Cas12Lambda2 | TATTTTGTATGGAGTAAACAAC |
| | Racr VA1 | TAAATTTCTACTGTTTGTAGAT |
| | Racr VA2 | TAAATTTCTACTGTTTGTAGAT |
| | Racr VA3 | TAAATTTCTACTATTTGTAGAT |
| | Cas12a2 | TAATTTCTACTGTTGTAGAT |
| | Cas12c2 | AGCAGGATTCAGGTTGGGTTTGAGG |
| | Cas12k | AGGTGGGTTGAAAG |
| | Cas12g | GGTGGAAAGGGCCGGAGATTTACCGGCTCTGACACC |

| | | |
|---|---|---|
| | Cas12L | GAAAAACGCTCTTAGGGAATGAAAG |
| | Cas12m | GTGTCATAGCCCAGCTTGGCGGGCGAAGGCCAAGAC |
| Cas13 | Cas13a | GGATTTAGACCACCCCAAAAATGAAGGGGACTAAAACA |
| | Cas13b | GTTGTGGAAGGTCCAGTTTTGAGGGGCTATTACAAC |
| | Cas13c | GACTAAAACCAAGTAAATTGGTATTTAAAC |
| | Cas13d | AACCCCTACCAACTGGTCGGGGTTTGAAAC |
| Cas9 | Cas9a | GTTTTAGAGCTATGCTGTTTTGAATGGTCCCAAAAC |
| | Cas9b | GTTTCAGTTGCTGAATTATTTGGTAAACT |
| | Cas9c | GTTGTAGCTCCCATTCTCATTTCG |
| | Cas9d | GTTACAGTTAAGGCTCT |
| Scramble | Poly A | AAAAAAAAAAAAAAAAAAAAAA |
| | Poly T | TTTTTTTTTTTTTTTTTTTTTT |
| | Poly C | CCCCCCCCCCCCCCCCCCCCCC |
| | AT | ATATATATATATATATATATAT |
| | GC | GCGCGCGCGCGCGCGCGCGCGC |
| | AC | ACACACACACACACACACACAC |
| | CT | CTCTCTCTCTCTCTCTCTCTCT |
| Spacer sequence | | ACAGCCCATCGACTGGTGTT |

**Table S8.** Primers and ΨDNA used for viral RNA and HCV patient sample detection

| Name | Sequence |
|---|---|
| DENV Geneblock w/ T7 [44] | AGTACATATTCAGGGGCCAACCTCTCAACAATGACGAAGACCATGCTC ACTGGACAGAAGCAAAAATGCTGCTGGACAACATCAACACACCAGAA GGGATTATACCAGCTCTCTTTGAACCAGAAAGGGAGAAGTCAGCCGCC ATAGACGGTGAATACCGCCTGAAGGGT |
| ZIKV Geneblock w/ T7 [44] | GACACCGGAACTCCACACTGGAACAACAAAGAAGCACTGGTAGAGTT CAAGGACGCACATGCCAAAAGGCAAACTGTCGTGGTTCTAGGGAGTC AAGAAGGAGCAGTTCACACGGCCCTTGCTGGAGCTCTGGAGGCTGAG ATGGATGGTGCAAAGGGAAGGCTGTCCTCTGGC |
| HCV-1a Geneblock w/ T7 | AGACACACTCCAGTCAATTCCTGGCTAGGCAACATAATCATGTTTGCC CCCACACTGTGGGCGAGGATGATACTGATGACCCACTTCTTTAGCGTC CTCATAGCCAGGGATCAGCTTGAACAGGCCCTTGATTGCGAGATCTAC GGAGCCTGCTACTCCATAGAACCACTGGATCTACCTCCAATCATTCAA AGACTCCATGGCCTCAGCGCGTTTTCACTCCACAGTTACTCTCCAGGTG AA |
| HCV-1b Geneblock w/ T7 | CAGCTAGACACACTCCAGTCAACTCCTGGCTAGGCAACATCATCATGT ATGCGCCCACCTTATGGGCAAGGATGATTCTGATGACTCACTTCTTCTC CATCCTTCTAGCTCAGGAGCAACTTGAAAAAGCCCTAGATTGTCAGAT CTACGGGGCCTGTTACTCCATTGAGCCACTTGACCTACCTCAGATCATT CAGCGACTCCATGGTCTTAGCGCATTTTCACTCCATAGTTACTCTCCAG GTGAG |
| DENV For Primer with T7 [44] | GAAATTAATACGACTCACTATAGGGGTACATATTCAGGGGCCAACCTC TC |
| DENV Rev Primer [44] | TTTCTGGTTCAAAGAGAGCTGGTAT |
| DENV AsCas12a ΨDNA | TCC AGT GAG CAT GGT CTT CGT AAT TTC TAC TAA GTG TAG AT |
| ZIKV For Primer with T7 [44] | GAAATTAATACGACTCACTATAGGG CCACACTGGAACAACAAAGAAGCAC |
| ZIKV Rev Primer [44] | ACAGCCTTCCCTTTGCACCATCCATCTCAG |
| ZIKV AsCas12a ΨDNA | GAA CCA CGA CAG TTT GCC TTT AAT TTC TAC TAA GTG TAG AT |
| HCV NS5B For Primer with T7 | GAAATTAATACGACTCACTATAGGGCACACTCCAGTCAATTCCTGGCT AGG |
| HCV NS5B Rev Primer | CACCTGGAGAGTAACTGTGGAGTGAA |
| HCV NS5B 1a AsCas12a ΨDNA | CAA GGG CCT GTT CAA GCT GAT AAT TTC TAC TAA GTG TAG AT |
| HCV NS5B 1b AsCas12a ΨDNA | CTA GGG CTT TTT CAA GTT GCT AAT TTC TAC TAA GTG TAG AT |
| HCV 5UTR For Primer with T7 | TAATACGACTCACTATAGGAGCGTCTAGCCATGGCGTT |
| HCV 5UTR Rev Primer | GCAAGCACCCTATCAGGCAGT |
| HCV 5UTR AsCas12a ΨDNA | TCCAAGAAAGGACCCGGTCGTAATTTCTACTAAGTGTAGAT |
| HCV E2 T7 For Primer with T7 | TAATACGACTCACTATAGACACCAACGGCAGTTGGCAC |
| HCV E2 Rev Primer | AAGGTGTTGTTGCCCACCCC |
| HCV E2 AsCas12a ΨDNA | GCCTGAAGAGTTGAATTTGTTAATTTCTACTAAGTGTAGAT |

**Table S9.** ΨDNA sequences for mCherry gene silecing and qPCR primers.

| Name | Sequence |
| --- | --- |
| ΨDNA 1<br>(targets mCherry start codon) | G*A*T*GATGGCCATGTTATCCTTAGATGTGAATCATCTTT*A*A*T |
| ΨDNA 1+<br>(targets mCherry start codon) | +G*+A*+T*GATGGCCATGTTAT+C+C+TTAGATGTGAATCATCTTT*A*A*T |
| ΨDNA 2<br>(targets mCherry) | T*T*G*GAGCCGTACATGAACTGTAGATGTGAATCATCTTT*A*A*T |
| Scramble DNA | TCGTCGGCAGCGTCTAATACGACTCACTATAGGG |
| qPCR mCherry For | GACTACTTGAAGCTGTCCTTCC |
| qPCR mCherry Rev | CGCAGCTTCACCTTGTAGAT |
| qPCR mCherry FAM Probe | FAM/TTCAAGTGG/ZEN/GAGCGCGTGATGAA/Iowa Black |
| qPCR GAPDH For [42] | GCTCCCTCTTTCTTTGCAGCAAT |
| qPCR GAPDH Rev [42] | TACCATGAGTCCTTCCACGATAC |
| qPCR GAPDH Cy5 Probe [42] | Cy5/TCCTGCACC/TAO/ACCAACTGCTTAGCACC/Iowa Black RQ |

*signifies phosphorothioated bonds
+indicate LNAs

**Table S10.** ΨDNA sequences for endogenous gene silecing and qPCR primers.

| Name | Sequence |
|---|---|
| ΨDNA NT derived from the crRNA in [9] | T*C*A*CCAGAAGCGTACCATACTCACGAACAGTAGATGTGAATCATCTTT*A*A*T |
| ΨDNA PPIA derived from the crRNA in [45] | A*A*A*CACCACATGCTTGCCATCCAACCACTCTAGATGTGAATCATCTTT*A*A*T |
| ΨDNA RPL4 derived from the crRNA in [45] | G*A*A*GTTCAGGAACTTCCTCAATACGATGACTAGATGTGAATCATCTTT*A*A*T |
| ΨDNA NRAS derived from the crRNA in [46] | C*T*G*GTCTTGGCTGAGGTTTCTAGATGTGAATCATCTTT*A*A*T |
| ΨDNA SMARCA4 derived from the crRNA in [46] | C*G*A*TGCGGTGGGCTCGGTCCTAGATGTGAATCATCTTT*A*A*T |
| ΨDNA PCSK9 #1 | A*A*G*CCAGGAAGAAGGCCATGGAAGACATGCTAGATGTGAATCATCTTT*A*A*T |
| ΨDNA PCSK9 #2 | A*T*G*GGGCAACTTCAAGGCCAGCTCCAGCAGTAGATGTGAATCATCTTT*A*A*T |
| ΨDNA PCSK9 #3 (Fig. 6) | C*C*A*GGTTCCACGGGATGCTCTGGGCAAAGATAGATGTGAATCATCTTT*A*A*T |
| ΨDNA PCSK9 #4 | G*G*A*AGACATGCAGGATCTTGGTGAGGTATCTAGATGTGAATCATCTTT*A*A*T |
| ΨDNA PCSK9 #5 | C*T*C*TGACTGCGAGAGGTGGGTCTCCTCCTTTAGATGTGAATCATCTTT*A*A*T |
| qPCR PPIA | Hs99999904_m1 |
| qPCR RPL4 | Hs00973287_g1 |
| qPCR PCSK9 | Hs00545399_m1 |
| qPCR NRAS | Hs00180035_m1 |
| qPCR SMARCA4 | Hs00231324_m1 |

*signifies phosphorothioated bonds
